# PAM-free diagnostics with diverse type V CRISPR-Cas systems

**DOI:** 10.1101/2024.05.02.24306194

**Authors:** Santosh R. Rananaware, Katelyn S. Meister, Grace M. Shoemaker, Emma K. Vesco, Luke Samuel W. Sandoval, Jordan G. Lewis, August P. Bodin, Vedant N. Karalkar, Ian H. Lange, Brianna Lauren Maria Pizzano, Minji Chang, M. Reza Ahmadimashhadi, Sarah J. Flannery, Long. T. Nguyen, Gary P. Wang, Piyush K. Jain

## Abstract

Type V CRISPR-Cas effectors have revolutionized molecular diagnostics by facilitating the detection of nucleic acid biomarkers. However, their dependence on the presence of protospacer adjacent motif (PAM) sites on the target double-stranded DNA (dsDNA) greatly limits their flexibility as diagnostic tools. Here we present a novel method named PICNIC that solves the PAM problem for CRISPR-based diagnostics with just a simple ∼10-min modification to contemporary CRISPR-detection protocols. Our method involves the separation of dsDNA into individual single-stranded DNA (ssDNA) strands through a high-temperature and high-pH treatment. We then detect the released ssDNA strands with diverse Cas12 enzymes in a PAM-free manner. We show the utility of PICNIC by successfully applying it for PAM-free detection with three different subtypes of the Cas12 family- Cas12a, Cas12b, and Cas12i. Notably, by combining PICNIC with a truncated 15-nucleotide spacer containing crRNA, we demonstrate PAM-independent detection of clinically important single-nucleotide polymorphisms with CRISPR. We apply this approach to detect the presence of a drug-resistant variant of HIV-1, specifically the K103N mutant, that lacks a PAM site in the vicinity of the mutation. Additionally, we successfully translate our approach to clinical samples by detecting and genotyping HCV-1a and HCV-1b variants with 100% specificity at a PAM-less site within the HCV genome. In summary, PICNIC is a simple yet groundbreaking method that enhances the flexibility and precision of CRISPR-Cas12-based diagnostics by eliminating the restriction of the PAM sequence.

## Introduction

The twenty-first century has experienced recurrent waves of global disease outbreaks, underscoring the need for rapid, high-throughput diagnostic tests^1,2^. Simultaneously, significant advances in Clustered Regularly Interspaced Short Palindromic Repeats (CRISPR)- based technology have presented a novel opportunity for molecular diagnostics through the detection of pathogenic nucleic acid signatures^3–8^. CRISPR originated from the prokaryotic adaptive immune system and is embedded in the genome of many bacteria and archaea^9–11^. The diversity of CRISPR-associated (Cas) enzymes offers many avenues for genetic exploration. Specifically, the class 2 type V CRISPR-Cas12 enzymes possess the capability of non-specific collateral cleavage or ‘trans-cleavage’ of ssDNA reporter molecules after target-specific cis-cleavage^12,13^.

Harnessing the trans-cleavage capabilities of Cas12 systems in conjunction with fluorescent ssDNA molecules can potentially streamline diagnostic processes by making them more accessible for point-of-care applications. CRISPR-based platforms such as SHERLOCK, DETECTR, SPADE, and ENHANCE have already been developed for portable, point-of-care diagnostics^5–8^ Nevertheless, a shared limitation among all these technologies is their strict dependence on the presence of a Protospacer Adjacent Motif (PAM) site, a short 2-5 nucleotide sequence that is located on the non-target strand of the DNA^14,15^. For example for CRISPR- Cas12a systems a short PAM motif consisting of a ’TTTV’ sequence is a pre-requisite for the binding and cleavage of the target by the Cas^16,17^. However, assuming a random and uniform distribution of nucleotides within a genome, the probability of finding a sequence like TTTV at any given position is merely 1.17%. This limitation poses a significant disadvantage as non-canonical PAM sequences either go entirely undetected or are sub-optimal in their performance, making CRISPR designs less programmable^3,18,19^. This is especially problematic when developing tests for mutational variants. Certain polymorphisms and single-point mutations that are identifiers for drug resistance and tumor development might not always be adjacent to a PAM site to enable detection^20–23^. Therefore, it is crucial to develop a versatile PAM-free CRISPR-Cas diagnostic platform.

While there are many efforts to loosen specific PAM requirements^16,24–29^ and an ongoing search to identify^30–35^ or engineer^16,36–40^ PAM-less variants, most lack simplicity, require multiple steps for amplification, and aren’t versatile enough to be used with other Cas enzymes. Hence, it is very desirable to develop an easier method with fewer steps and fewer constraints. To overcome this limitation, here we present a novel method named PICNIC which stands for <u>P</u>AM-less Identifi<u>c</u>ation of <u>N</u>ucleic Acids with <u>C</u>RISPR. PICNIC achieves a truly PAM-free detection with different types of Cas12 enzymes with just a simple ∼10-min modification to contemporary CRISPR-detection protocols and requires only commonly available lab reagents to work.

The basis of PICNIC lies in the previously reported observation that the stringent PAM requirement for CRISPR-Cas systems is only found when detecting double-stranded DNA (dsDNA) but not single-stranded DNA (ssDNA) ^12^. In the case of dsDNA, PAM recognition initiates the unwinding of the duplex, opening the target region for hybridization^15,30^. For ssDNA substrates, this unwinding step is not required, thus the PAM constraint is bypassed. We hypothesized that separating the target dsDNA into separate ssDNA strands through a combination of high heat and pH may destabilize the target and enable PAM-free detection for a multitude of Cas enzymes.

In this work, we have developed and successfully applied PICNIC for the detection of a library of non-canonical PAM sequences using several different type V Cas enzymes, including Cas12a^41,42^, Cas12b^43,44^, and Cas12i^45^. PICNIC works essentially by subjecting the target DNA to a high temperature and high pH treatment for 2-10 min before detection with CRISPR. The key idea here is that while the high temperature initially disrupts the dsDNA duplex, the highly basic pH environment prevents the separated strands from re-annealing even when the temperature is subsequently lowered to enable CRISPR-based detection. This simple modification to the traditional diagnostic protocol separates the target dsDNA into single strands and enables the detection of virtually any PAM sequence, thus providing complete freedom over the choice of target site.

We observed that PICNIC is not only able to detect any non-canonical PAM sequence with high accuracy but in the case of Cas12b and Cas12i, was also able to increase the overall fluorescence signal obtained from trans-cleavage of reporters. Furthermore, by combining PICNIC with a guide RNA containing a short 15-nt spacer sequence we were able to discriminate the K103N single nucleotide polymorphisms (SNP) in synthetic Human Immunodeficiency Virus (HIV) cDNA, revealing the potential for identifying drug-resistant strains of HIV with PICNIC. This is important because clinically important SNPs might not always have a nearby PAM sequence available, thus making their detection through contemporary methods difficult. However, we show that SNP discrimination at a PAM-less site can be reliably accomplished with PICNIC. Finally, we utilized the versatility of PICNIC for detecting and genotyping HCV variants such as HCV-1a and HCV-1b at a highly mutated region within the HCV genome, albeit with no available PAM site. Using our approach, we were able to detect both variants with 100% specificity and 100% (HCV-1a) and 95.4% (HCV- 1b) accuracy respectively. Thus, our work here provides a simple yet remarkably powerful solution to the PAM problem in CRISPR diagnostics.

## Results

### Development of PICNIC method for PAM-free detection

The class 2 type-V CRISPR-Cas enzyme Cas12a is known to require a short T-rich PAM sequence to initiate target recognition and cleavage. However, the PAM sequence is known to be a requirement for double-stranded DNA (dsDNA) but not single-stranded DNA (ssDNA) substrates. We hypothesized that converting the dsDNA to ssDNA with heat and pH might enable PAM-free detection with CRISPR-Cas12a **(Fig. 1a)**.

**Figure 1:**
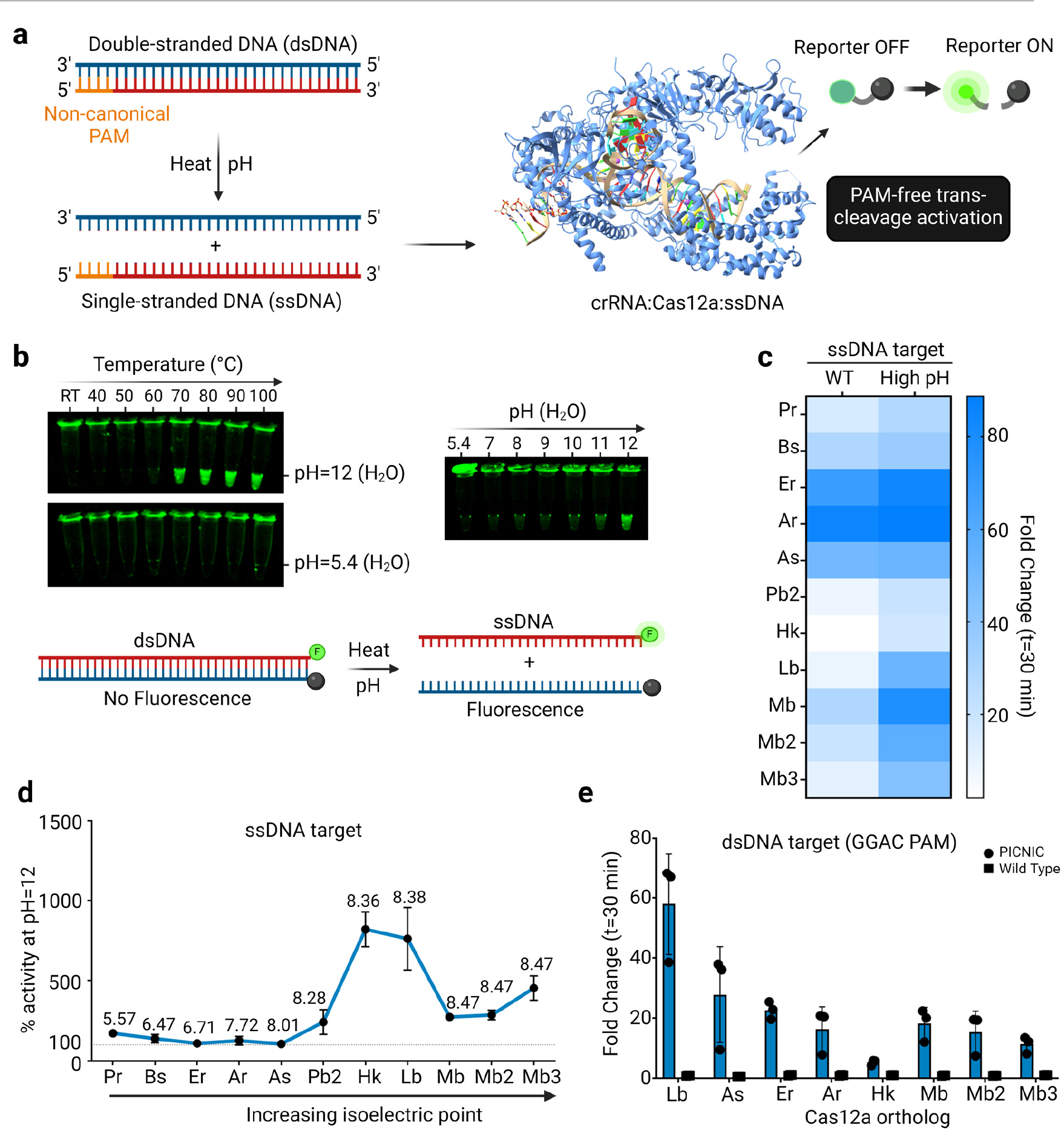
DNA denaturation dynamics and establishment of PICNIC method for PAM-free detection (a) Schematic of PICNIC method for PAM-free detection. A double-stranded DNA (dsDNA) target containing any non-canonical PAM sequence can be denatured to single-stranded DNA (ssDNA) strands using a combination of high pH and temperature. Each ssDNA strand can then be detected in a PAM-free manner using the target-dependent trans-cleavage activity of CRISPR-Cas12 systems. Crystal structure of LbCas12a for representation was obtained from PDB: 5XUS ^31^ **(b)** The effect of temperature and pH on the stability of dsDNA is tested using a FAM and quencher labeled target. The target shows no fluorescence in dsDNA form due to the proximity of the FAM and quencher but fluoresces when denatured to ssDNA. The target was heated at various temperatures in regular nuclease-free water or pH 12 water for 10-min and then cooled down (left) or heated at 100 °C in different pH conditions and then cooled down (right). Fluorescence in the cooled samples was visualized under blue light (Ex 485/20, Em 528/20). **(c)** Detection of a ssDNA target using 11 different Cas12a orthologs with a conventional CRISPR detection (WT) or detection with a modified method where the target is heated to 85°C in a pH 12 buffer before detection (High pH). Heat map represents the fold change in fluorescence intensity as compared to a no target control (NTC) at time t = 30 min. **(d)** Plot representing the % increase in trans-cleavage activity with the high pH method as compared to the conventional WT method for 11 different Cas12a orthologs. **(e)** A dsDNA target containing a non-canonical GGAG PAM was denatured by heating at 85°C for 10 min in PICNIC buffer and was detected using the trans-cleavage assay with 8 different orthologs of CRISPR-Cas12a. Fold changes in fluorescence intensities relative to the No Target Control (NTC) at t = 60 min are shown. Mean ± SD (n = 3) are indicated.

To explore the effect of temperature and pH on the stability of dsDNA, we conducted a series of experiments using a target dsDNA labeled with a fluorophore (FAM) on one strand and a quencher on the complementary strand. In the dsDNA form, this target exhibits no fluorescence due to the quenching. However, when denatured to ssDNA, fluorescence is observable. To test the effect of heating on the integrity of the dsDNA target, we first heated it under various temperatures ranging from room temperature (rt) to 100°C in either nuclease-free water (pH 5.4) or a pH-adjusted water (pH 12) for 10-min, followed by a 15-min cooling down phase **(Fig. 1b, left)**. Concurrently, we also subjected the targets to heating at 100°C under different pH, ranging from 7 to 12, followed by cooling **(Fig. 1b, right)**. In both cases, fluorescence in the cooled samples was visualized under blue light.

Our results indicated that when the target is heated in regular nuclease-free water (pH 5.4) and then cooled back down, it goes back to being double-stranded irrespective of the temperature that it is initially heated at. However, in the basic water (pH 12), we observed that when heated at or above 70°C, the target exhibited a high level of fluorescence even after cooling down, suggesting that it remains single-stranded despite being cooled down, likely due to the high pH of the solution inhibiting the hydrogen bonding and subsequent re-annealing of the separated DNA strands **(Fig. S1)**. Our observations with the target heating and cooling under different pH conditions confirmed this hypothesis as we observed increased fluorescence in the samples with increasing basicity of the solution, with pH of 12 displaying the highest levels of fluorescence **(Fig. S2)**.

To test the tolerance of Cas12a orthologs towards a highly basic pH environment, we tested the detection of a ssDNA target with 11 different Cas12a orthologs with either a contemporary wild-type CRISPR/Cas-based detection method (WT) or a modified protocol where the ssDNA target was heated to 85°C at pH of 12 for 10-min before transferring it to a CRISPR/Cas detection reaction consisting of the target-specific crRNA, Cas12a, NEB2.1^TM^ buffer (final pH = 8.3), and fluorophore-quencher (FQ) based DNA reporter molecules. We observed that all the Cas12a orthologs were able to tolerate the higher reaction pH and showed an identical or improved detection towards the ssDNA target in the high pH method compared to the WT (**Fig. 1c).** Interestingly, Cas12a enzymes with isoelectric points (pI) above 8.3 showed improved activity in the modified method as compared to the WT suggesting that the high pH reaction mix might be better tolerated by enzymes having a higher pI **(Fig. 1d)**. The greatest improvement was observed for LbCas12a and HkCas12a orthologs.

To test if our method utilizing a high pH can be used to detect PAM-less dsDNA substrates, we designed a target containing a non-canonical GGAG PAM flanking the target sequence. We first made this target single-stranded by subjecting it to heat-denaturation at a high pH (85°C, pH 12) and then cooled it down for use in the CRISPR-Cas12a trans-cleavage reaction (final pH = 8.3). We tested this target with 8 different orthologs of Cas12a that showed a high trans-cleavage activity in the previous experiment and that span a wide range of pI within the Cas12a-family **(Fig. 1e)**. All 8 enzymes detected the non-canonical PAM-containing target, thereby validating our hypothesis of PAM-free detection through this method. Among the enzymes tested, LbCas12a (pI 8.38) exhibited the highest activity. We name our method of subjecting the dsDNA to denaturation by high temperature and pH as PICNIC. Thus, for the remainder of this work, the term ’PICNIC-step’ refers to the heating of the target DNA sample at a high temperature (typically 85°C for 10 min, unless specified otherwise) in a highly basic buffer (typically water at pH of 12, unless specified otherwise) done before the CRISPR-step.

### PAM-free detection with CRISPR-Cas12a

To optimize the PICNIC step for different incubation conditions, we tested the heating of the non-canonical GGAG PAM-containing target under different pH, temperature, and incubation times, and then tested its detection with LbCas12a to identify the best conditions. Upon heating it in a pH 12 buffer for 10-min at different incubation temperatures spanning rt to 95°C before cooling down, we observed the highest trans-cleavage activity for incubation temperatures of 85°C and 95°C **(Fig. S3)**. Temperatures lower than 85°C failed to induce any trans-cleavage activity, suggesting that higher temperatures are better for PICNIC. Next, we incubated the target at 85°C and pH 12 buffer for different incubation times ranging from 0 min to 30 min before cooling down, we observed the greatest level of detection for a 10-min incubation step (**Fig. S4**). Furthermore, upon heating the target at different pH conditions spanning between 7 to 12 before cooling down, we observed the highest detection at pH of 12 **(Fig. S5)**. We also tested different types of additives to the PICNIC buffer and observed that adding a small concentration of Tween-20® detergent to the buffer had a slight increase in the overall activity **(Fig. S6)**. Thus, the optimized heating conditions for the PICNIC step were finalized to be pH 12, 85°C for a 10-minute incubation step with the addition of 0.5% Tween-20® detergent in the buffer.

To demonstrate that the optimized PICNIC method can detect any non-canonical PAM presented, we designed a library of 64 unique PAM containing double-stranded DNA activators containing NNNG PAM adjacent to the target site **(Fig. 2a)**. We then characterized the detection of this PAM library using either the WT CRISPR-Cas12a **(Fig. 2b)** or PICNIC **(Fig. 2c).** Interestingly, we observed that even with the WT CRISPR-Cas12a, target sequences containing a cytosine (C) or a thymine (T) nucleotide at the 2^nd^ position were detected equally well, suggesting that the PAM requirement for LbCas12a for *in vitro* trans-cleavage is different as compared to the TTTV PAM that has been previously reported inside cells. The tolerance of these non-canonical PAM sequences by Cas12a orthologs has also been previously reported by other papers^31,32^. Nevertheless, PAM sequences containing adenine (A) or guanine (G) nucleotides at the 2^nd^ position could not be detected with the WT LbCas12a. Remarkably, PICNIC was able to detect all 64 combinations of PAM sequences, thus demonstrating its capability as a true PAM-less method (**Fig. 2c).**

**Figure 2:**
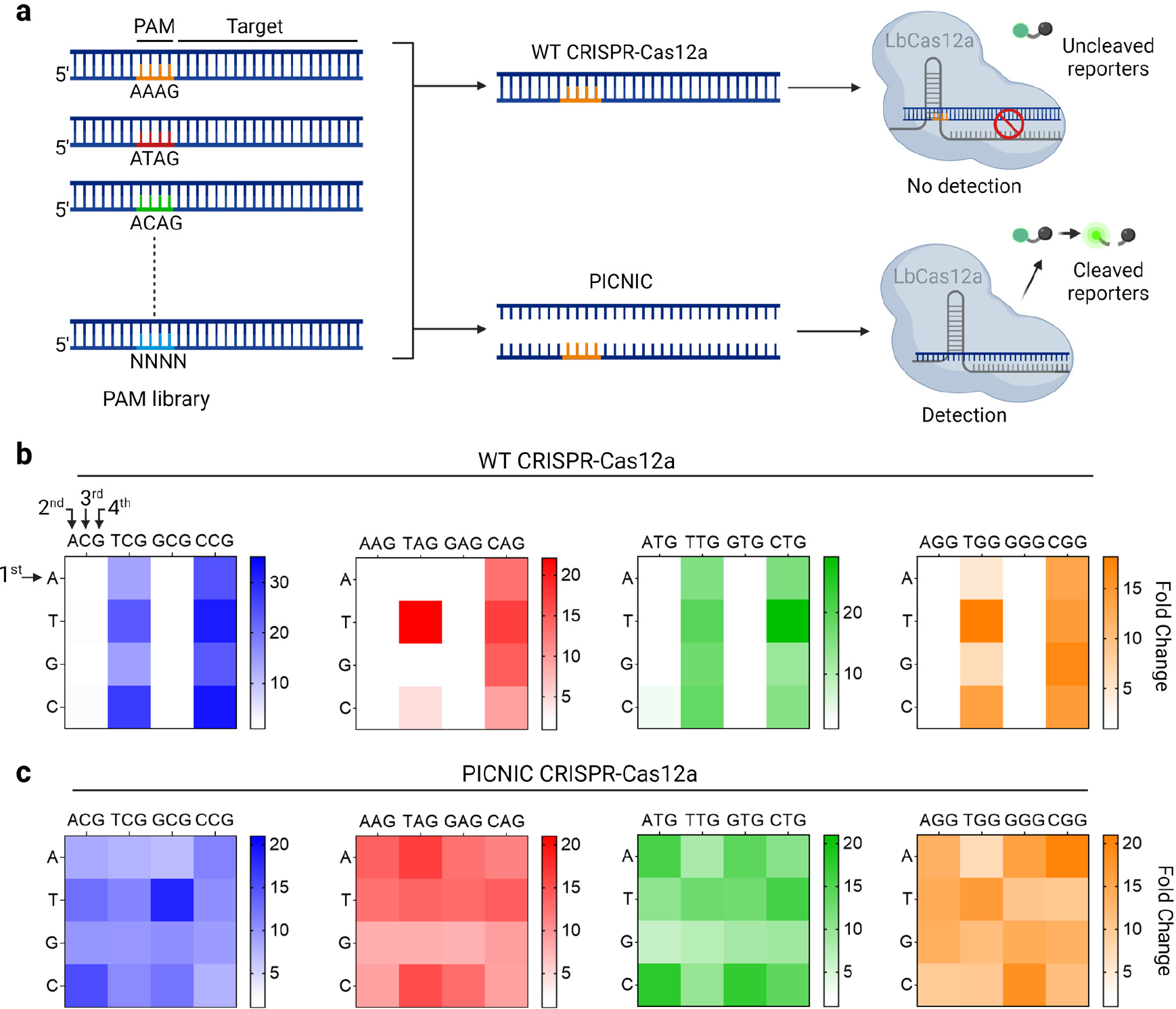
Comparison of PAM sequence detection between WT CRISPR-Cas12a and PICNIC (a) Schematic detailing the detection of a library of dsDNA targets, each containing a distinct PAM sequence, with WT CRISPR-Cas12a or the PICNIC method. A dsDNA target without the correct PAM sequence cannot initiate trans-cleavage activity of WT CRISPR-Cas12a but can do so only if the PICNIC method is applied. **(b-c)** Detection of a library of dsDNA activators with different PAM sequences with either the WT CRISPR-Cas12a (b) or PICNIC (c). Heat map represents the fold-change of fluorescence intensity relative to NTC at time t = 30 min (n = 3). Each cell within the heat map represents the detection of a unique PAM sequence which can be read from the 5’-end to the 3’-end with nucleotides labeled as 1^st^, 2^nd^, 3^rd,^ or 4^th^ in the figure.

For the originally designed PAM library, we fixed the 4^th^ nucleotide to be a constant ’G’ across the different PAM sequences in order to reduce the order of the PAM library from 256 to 64. To study the effect of the 4^th^ position of the PAM on the trans-cleavage activity by WT CRISPR-Cas12a as well as PICNIC, we designed another set of activators, where the first 3 nucleotides were fixed to be either AAA, TTT, GGG, or CCC while the 4^th^ nucleotide was varied to be A, T, G, or C. We then tested these new targets with both the WT CRISPR as well as PICNIC. Similar to the previous results, we observed that the WT-CRISPR only detected PAM sequences containing a ‘C’ or a ‘T’ at the 2^nd^ position, while with the PICNIC method, we were able to detect every single PAM sequence **(Fig. S7)**.

### PICNIC can be expanded to diverse type-V CRISPR-Cas systems

Next, we wondered whether our PICNIC method could be expanded to other type-V CRISPR-Cas systems that also need a PAM. For this purpose, we selected BrCas12b (pI 9.36) and Cas12i1 (pI 9.28) on account of their relatively high isoelectric points. BrCas12b is classified as a type V-B Cas protein derived from *Brevibacillus sp.* SYP-B805 and it has been empirically determined that the canonical PAM is TTN^8^ **(Fig. 3a)**.

**Figure 3:**
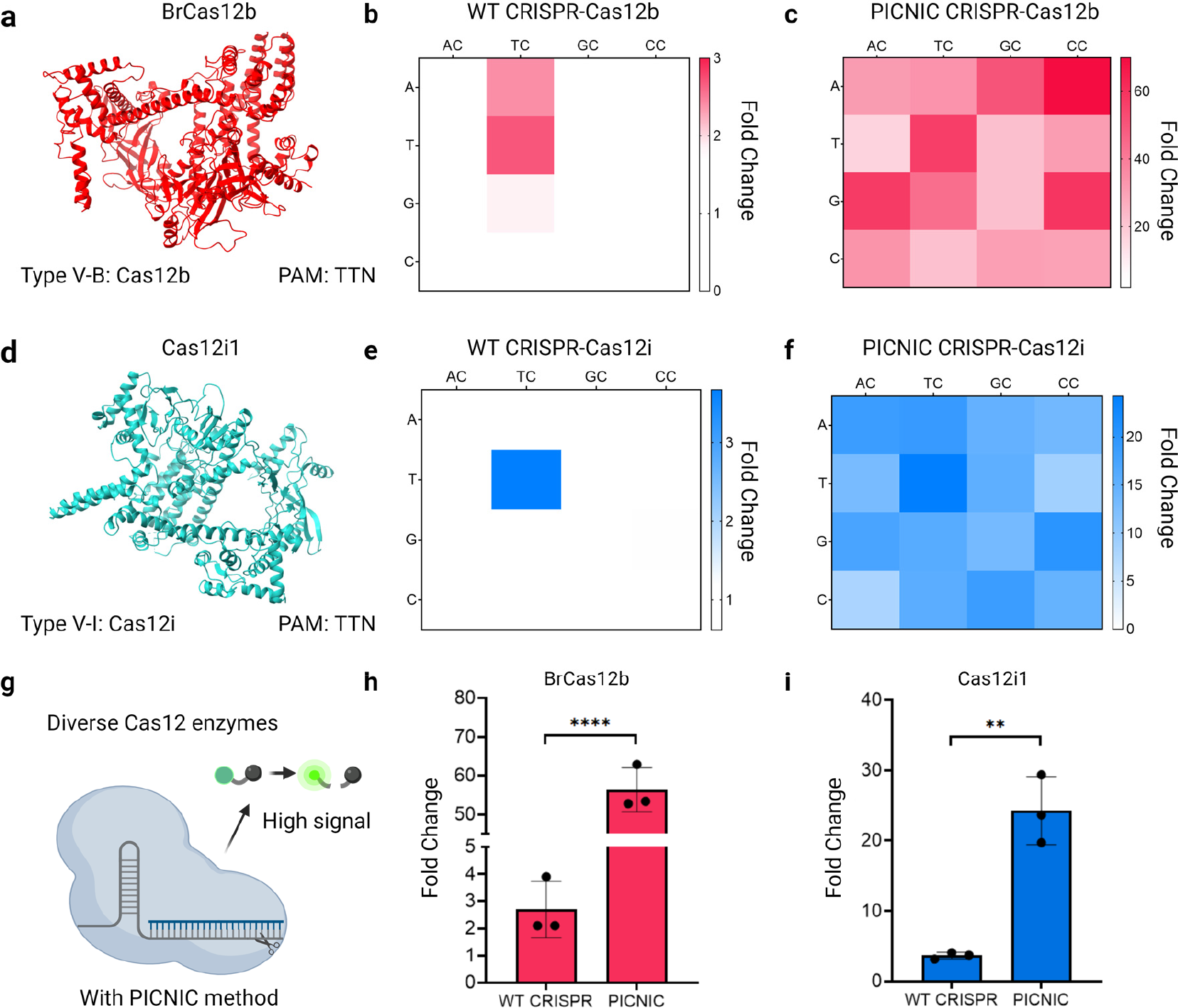
PAM-free detection with CRISPR-Cas12b and Cas12i (a) Structure for BrCas12b predicted using Alphafold^58^. **(b-c)** Detection of a library of dsDNA activators with different PAM sequences with either the (b) WT CRISPR-Cas12b or (c) PICNIC. **(d)** The crystal structure for Cas12i1 was obtained from PDB: 7D8C^59^. **(e-f)** Detection of a library of dsDNA activators with different PAM sequences with either the (e) WT CRISPR-Cas12i or (f) PICNIC. **(g)** PICNIC method boosts the signal generated from trans-cleavage of targets containing canonical or non-canonical PAM by diverse Cas12 enzymes **(h-i)** Comparison of the fold change in fluorescence intensity for detection of the canonical ‘TTC’ PAM with WT-CRISPR and PICNIC using (h) BrCas12b or (i) Cas12i1**. (b-c, e-f)** Heat map represents the fold-change of fluorescence intensity relative to NTC at time t = 30 min (n = 3). Each cell within the heat map represents the detection of a unique PAM sequence which can be read from the 5’- end to the 3’-end with nucleotides labeled as 1^st^, 2^nd^, or 3^rd,^ in the figure. Fold changes in fluorescence intensities relative to the No Target Control (NTC) at t = 60 min are shown. Mean ± SD (n = 3) are indicated. Statistical analysis was performed using a two-tailed t-test where ns = not significant with p > 0.05, and the asterisks (* P ≤ 0.05, ** P ≤ 0.01, *** P ≤ 0.001, **** P ≤ 0.0001) denote significant differences.

More specifically, for this study, we chose to use the engineered variant of BrCas12b- RFND that we have previously reported. We found that this variant increases the thermostability and enhances the trans-cleavage activity as compared to the WT BrCas12b^46^.

We first characterized the PAM requirement for the trans-cleavage activity for this enzyme using a subset of the PAM library used in the previous figure. Upon testing the wild-type activity on the PAM library we observed BrCas12b to strongly tolerate the TTC and ATC PAMs while a weak but noticeable activity with the GTC PAM **(Fig. 3b)**. When the same PAM library was tested using the PICNIC method, we observed that all possible PAM combinations were detected **(Fig. 3c, S8)**.

Similarly, for Cas12i, we used the originally identified Cas12i1 ortholog, which is reported to have a high level of trans-cleavage activity^47^ **(Fig. 3d)**. Despite its tremendous potential, currently, there are no reports of harnessing the trans-cleavage activity of Cas12i for diagnostic purposes as has been done previously for Cas12a, Cas12b, and Cas12c. Cas12i1 had trans-cleavage activity for the canonical TTC PAM as expected, no other PAM sequences were detected **(Fig. 3e)**. When tested with PICNIC, all variants of PAM were detected by Cas12i1, consistent with the results obtained for LbCas12a and BrCas12b **(Fig. 3f, S8)**

We also observed a significant increase in the overall detectable fluorescence levels when using PICNIC suggesting that these two enzymes have a much greater trans-cleavage activity for ssDNA as compared to dsDNA. Specifically, the activity of BrCas12b, as measured by the fold-change in the fluorescence intensity as compared to the NTC, increased from 3-fold with the WT-CRISPR to 60-fold with PICNIC. Similarly, the activity of Cas12i1 increased from 3- fold with WT-CRISPR to 20-fold with PICNIC **(Fig. 3 g-i)**. We hypothesized that these enzymes have a much higher preference for ssDNA than dsDNA as the substrate for initiating trans-cleavage, which might explain the increase in activity observed with PICNIC.

We also compared the trans-cleavage activities of LbCas12a, BrCas12b, and Cas12i1 with PICNIC for the detection of different concentrations of a non-canonical PAM-containing target. We observed that while all three enzymes are active for PAM-free detection with PICNIC, LbCas12a has the highest overall activity (**Fig. S9**). Thus, we demonstrate that PICNIC can be applied to multiple subtypes of Cas12 enzymes to convert them into a truly PAM-free diagnostic platform.

Finally, the thermostable nature of CRISPR-Cas12b has previously been shown to be useful for the development of one-pot CRISPR-based tests that are convenient to use on the field^46,48^. To validate that our approach can be translated to a one-pot reaction, we tested the detection of a library of PAM-variant targets in a one-pot reaction with BrCas12b. Specifically, the target was added to a one-pot mix containing reverse transcription loop-mediated isothermal amplification (RT-LAMP) primers, RT-LAMP reaction mix, guide RNA, BrCas12b, reporters, and PICNIC buffer. This reaction was heated at 62°C for 15-min and fluorescence was measured every 30s. Our results indicated that all the PAM sequences were successfully detected even in the one-pot reaction, thus, showing great potential for the use of PICNIC in a one-pot setting **(Fig. S10)**.

### PICNIC combined with truncated guide RNA enables SNP discrimination

The detection of SNPs is critical in various aspects of biomedical research and clinical diagnostics, particularly in the context of genetic disorders, infectious diseases, and personalized medicine. SNPs, which are variations of a single nucleotide in the genome, can have significant implications on an individual’s susceptibility to diseases, response to drugs, and other physiological traits. However, the precise detection of SNPs with CRISPR remains a challenge, especially in regions of the genome that lack accessible PAM sites.

Since we can overcome the PAM constraint using PICNIC, we wondered whether we could apply our method to the detection of clinically relevant SNPs. However, previous studies evaluating the activity of Cas12a have demonstrated that ssDNA-activators greatly tolerate single and dual base pair mismatches between the target and the crRNA^3^. To increase the specificity of PICNIC, which relies on ssDNA activation, we hypothesized that truncating the crRNA spacer sequence may lower or eliminate non-specific activation, thereby decreasing Cas12a’s mismatch tolerance and potentially enabling SNP discrimination.

To test this hypothesis, we systematically designed truncated crRNA of lengths 17-nt and 15- nt by trimming the bases of the full 20-nt spacer of the crRNA at the 3’-end. We tested the detection capabilities of the truncated crRNAs with target activators containing a non-canonical PAM and harboring 1 to 6 mutations near the seed region of the crRNA **(Fig. 4a)**. From this data, we found that the most sensitive difference in activity between single point mutations occurred between WT target and SNP (referred to as 1-mut in the figure) for the 15-nt guide and between the 4-mut and 5-mut target for the 20-nt guide **(Fig. 4b)**. This suggested to us that while a full guide can tolerate as many as 4 mutations in the target ssDNA, potentially a truncated guide RNA with a 15-nt spacer might be more suitable for detecting point mutations as compared to a canonical 20-nt spacer containing guide RNA.

**Figure 4:**
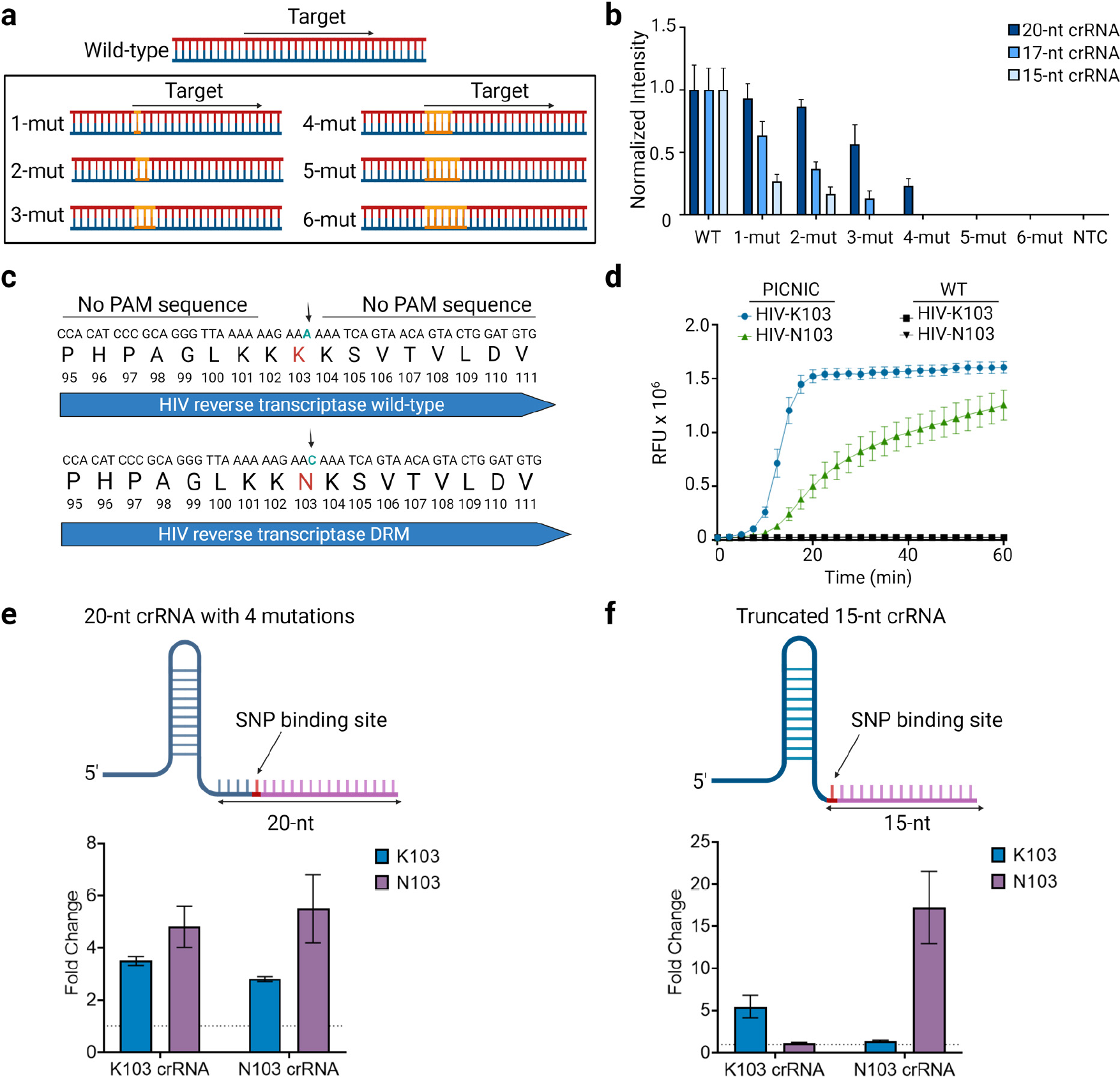
Detecting drug-resistant variants of HIV with PICNIC (a) Schematic of a dsDNA target sequence containing either wild-type reverse transcriptase (WT) or 1-6 mutations in the crRNA-binding region (1-mut to 6-mut). **(b)** Data representing detection of WT and 1-mut to 6-mut targets with crRNA containing spacers of lengths 20-nt, 17-nt, or 15-nt. This plot represents fluorescence intensity for each target normalized to the intensity of the WT target. Mean ± SD (n = 3) is indicated. **(c)** Schematic depicting the sequence of a portion of the human immunodeficiency virus (HIV) cDNA from amino acid positions 95-111 of the RT enzyme. The mutation K103N SNP from nucleobase A to nucleobase C, and is known to confer resistance against non-nucleoside reverse transcriptase inhibitors (NNRTI) targeting HIV. **(d)** Data depicting detection of HIV-K103 and HIV-N103 with the PICNIC method versus the WT Cas12a. The graph depicts the raw fluorescence units (RFU) for the respective complementary 20-nt crRNAs over 60 minutes **(e)** Schematic portraying the 20-nt crRNA, containing a randomized first 4 nucleotides followed by the SNP binding site at the 5th position of the spacer. Data representing SNP detection of K103 and N103 utilizing the respective 20-nt crRNAs is displayed below. **(f)** Schematic of the truncated 15-nt guide that contains SNP binding at the 1st position of the mutation site. SNP detection of K103 and N103 utilizing the respective 15-nt crRNAs is displayed below. **(e-f)** Fold change of fluorescence intensities relative to the NTC at t = 30 min are shown. **(b, e-f)** Mean ± SD (n = 3) are indicated.

We decided to apply our approach to the detection of the commonly occurring K103N mutation in the Human Immunodeficiency Virus (HIV) genome. This particular mutation is associated with high-level resistance to certain antiretroviral drugs, particularly the non-nucleoside reverse transcriptase inhibitors (NNRTIs) such as Nevirapine and Efavirenz^49,50^. Importantly, this mutation arises due to just a SNP, where an A or G at position 103 in the reverse transcriptase gene of HIV is changed to a C or T. This region contains no PAM sequences, making it undetectable with canonical PAM-restricted CRISPR methods **(Fig. 4c)**.

To validate that HIV-K103 and the drug-resistant mutant (DRM) HIV-N103 can both be detected using PICNIC, we designed two fully complementary 20-nt crRNAs for K103 and N103 that contain the canonical ‘A’ nucleotide (HIV-K103) or the mutation ‘C’ (HIV-N103) at the first position of the seed region. Both guides were subsequently tested to detect the cDNA of the WT as well as the DRM target **(Fig. 4d)**. Under PICNIC conditions, both K103 and N103 assays showed robust trans-cleavage activity. Unsurprisingly, both guides failed without PICNIC. This result is expected as this target region contains a non-canonical PAM sequence and thus, cannot be detected without PICNIC.

While the ability to detect a canonically difficult region using PICNIC is great, it is not sufficient. We also need to be able to discriminate between the WT and the DRM variant of HIV. Utilizing the results from Fig. 4b, we devised two strategies to enable SNP discrimination. First, we designed a 20-nt crRNA with 4 mutations in the seed region such that the SNP binds at the 5^th^ position. Our second strategy employed the use of a truncated 15-nt crRNA such that the SNP binds at the 1^st^ nucleotide within the crRNA spacer. Each guide was tested for SNP discrimination between synthetic cDNA targets containing either the K103 or N103 target using PICNIC **(Fig. 4e,f)**. The 20-nt guide containing four mutations did not show SNP discrimination for both the K103 as well as the N103 crRNAs. Conversely, the 15-nt guide exhibited a significant reduction in activity for the targets containing the SNP. Thus, robust SNP discrimination between PAMless targets (K103 and N103) can be attained with CRISPR- Cas12a by utilizing a truncated 15-nt guide and PICNIC.

To test if the strategy of using a truncated 15-nt guide can be applied universally to detect any possible single base mutations, we designed targets containing a single point A, T, G, or C mutation as well as a library of truncated crRNA containing A, U, G, or C at the first position of the spacer. Upon testing all the targets/crRNA in a combinatorial manner, we observed that truncated 15-nt crRNAs containing either G or C base at the first position can robustly discriminate between SNPs but crRNAs containing A or U base in the first position show non-specific activity for other mutations **(Fig. S11)**. Thus, our approach using 15-nt truncated guides for SNP discrimination worked well for SNPs containing a G or C base but not for A to T or T to A mutants.

### PICNIC can genotype HCV-1a and HCV-1b from human serum samples

Hepatitis C virus (HCV) infection is a leading cause of chronic liver disease worldwide, with significant variability in its clinical outcomes attributable in part to genetic diversity^51,52^. HCV genotyping is crucial in guiding direct-acting antiviral therapy, as the response to treatment can vary greatly depending on the specific HCV genotype. Genotypes 1a and 1b, in particular, are associated with different disease progression and are known for their varied response rates to direct-acting antiviral therapy and treatment outcomes^53^. Current guidelines recommend genotypes 1a and 1b be assessed before starting treatment. Therefore, precise, and rapid genotyping is imperative for effective clinical management of chronic HCV infection. However, genotyping HCV variants with traditional CRISPR-based diagnostics can become challenging due to the restrictions posed by the PAM sequence for guide RNA design.

We aligned the sequences of 6 major variants of HCV (1a, 1b, 1c, 2a, 2b, 3a) and identified a position near the non-structural-5B (ns5B) region within the HCV genome that is conserved within the population of each sub-group but has a high variability between the sub-groups. This region has previously been used for genotyping HCV variants with lab-based methods such as qPCR^54^, but the lack of a PAM site makes it difficult to detect with CRISPR. We designed specific crRNAs for each variant at this variable region such that the seed region of the crRNA matched with a 4-nt block that is variable between each HCV sub-type. The PAM sequences flanking these crRNA were non-canonical GGGA/AGGA/AAGA **(Fig. 5a).**

**Figure 5:**
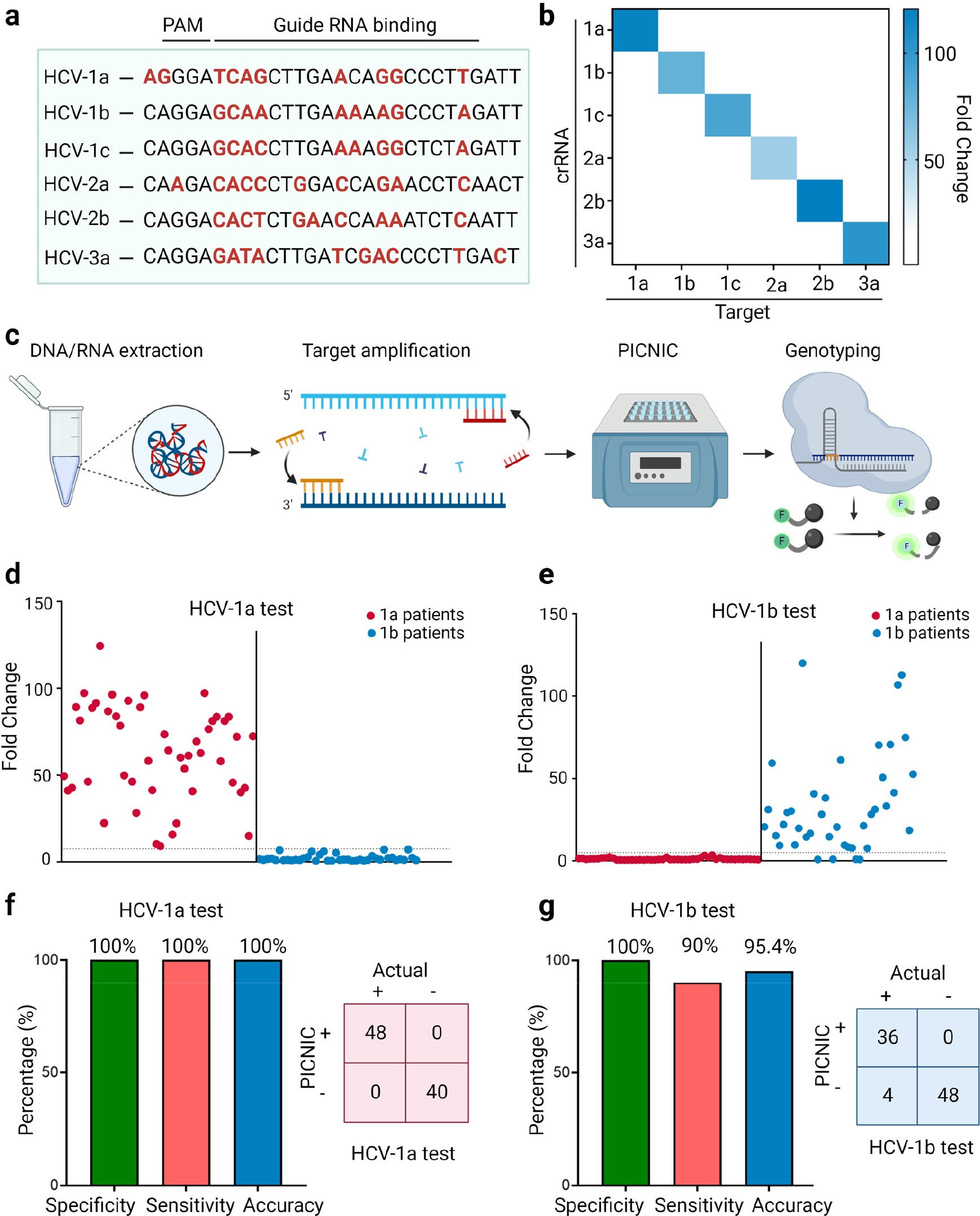
HCV variant genotyping with PICNIC (a) Schematic showing the alignment of 6 major variants of HCV with mutations relative to the consensus sequence highlighted in red. Specific guide RNAs for detecting and genotyping each variant are designed to target the area highlighted as ‘Guide RNA binding’. These guide RNAs target a non-canonical GGGA/AGGA/AAGA PAM. **(b)** Heat map depicting the specificity of crRNAs targeting synthetic cDNA mimics of HCV-1a, 1b, 1c, 2a, 2b, 3a towards their corresponding target at the non-canonical PAM region depicted in panel (a). The heat map represents the fold change in fluorescence intensity compared to the NTC for each crRNA/target combination after detection with PICNIC. The crRNA designed for each variant displays fluorescence only in the presence of its native target thus demonstrating good specificity. **(c)** Schematic showing the workflow for genotyping of HCV-1a and HCV-1b from human patient samples. The sample is first subjected to nucleic acid extraction, followed by target amplification with common primers. The amplified target is subjected to denaturation with the PICNIC method and subsequently processed with specific crRNA designed to genotype HCV-1a and HCV-1b. **(d-e)** Data represents detection and genotyping for 48 HCV-1a (red) and 40 HCV-1b (blue) samples from human serum using PICNIC. Panel c represents detection of the 88 samples with a crRNA specific for HCV-1a while panel d shows the same but with a crRNA specific for HCV-1b. The dot plot shows the fluorescence intensity for each patient sample as compared to the NTC. The dotted line indicates a detection threshold of 7.5-fold for the HCV-1a test and 5-fold for the HCV-1b test. **(f-g)** The plot represents the sensitivity, accuracy, and specificity for HCV-1a specific test (panel e) and HCV-1b test (panel f) respectively.

First, we evaluated the specificity of each crRNA to detect their respective target. For this purpose, we designed cDNA mimics of HCV ns5B for each of the 6 variants. We detected the 6 crRNAs and their corresponding targets in a combinatorial manner using PICNIC. We observed that each crRNA was able to detect its own target with a high sensitivity as well as a robust specificity **(Fig. 5b).**

Next, we decided to apply our test for genotyping clinical patient samples with PICNIC. To this end, we obtained 48 HCV-1a positive and 40 HCV-1b positive human serum samples. Patient serum samples were first processed to extract viral RNA, which was then reverse transcribed and amplified to get a high amount of cDNA **(Fig. 5c)**. PICNIC method with guides specific to variant 1a (HCV-1a test) and 1b (HCV-1b test) were used to detect and discriminate the patient samples **(Fig. 5 d,e)**. Remarkably, the HCV-1a test was able to discriminate the HCV-1a samples from the HCV-1b samples with 100% specificity as well as 100% sensitivity **(Fig. S12, 13)**. While the HCV-1b test had a 100% specificity in distinguishing the 1b samples from 1a, we observed 4 false negative results for the 1b samples bringing down the sensitivity to 90% for the HCV-1b test **(Fig. 5 f,g).** The false negative results can be attributed to either a low viral RNA load within these samples or a potential mutation/mismatch at this region between the crRNA and the target. Issues arising from mutation due to genomic variability in the HCV-1b variant can potentially be fixed by the usage of mixed bases within the crRNA. Thus, we establish that PICNIC can be utilized to successfully discriminate between HCV-1a and HCV-1b as well as potentially other HCV sub-types at a PAM-less region.

## Discussion

The development of rapid and accessible diagnostic platforms is imperative for tracking and controlling the spread of infectious diseases. Here, we have developed a novel method named PICNIC, which allows for a simple and PAM-free nucleic acid detection platform. The introduction of this technology addresses a critical limitation in conventional CRISPR-based diagnostics and, importantly, enables improved programmability to distinguish between different disease variants and monitor their spread. PICNIC fundamentally broadens the application of CRISPR-based diagnostic tools, which are typically constrained by the requirement of a PAM site for target detection. In conventional systems, the PAM requirement restricts the programmability of these platforms, making them less effective in performing operations such as distinguishing single-point mutations, detecting important markers for drug resistance as well as monitoring tumor development, and disease progression. By eliminating the PAM requirement, PICNIC allows for improved detection of these critical variations. PICNIC is a versatile, universal approach to do a truly PAM-free detection with a multitude of type V Cas enzyme sub-types such as Cas12a, Cas12b, Cas12i, and their orthologs. Importantly, this method is easy to use, fast (2-10 min), and only requires widely available lab reagents such as NaOH and a detergent like Tween-20 to make a highly basic buffer to break open the dsDNA. This opens up possibilities for a broader range of applications and provides the opportunity to choose the most suitable target site based on specific user requirements for the diagnostic assay without having to deal with the constraint of a PAM.

Another notable advantage of the PICNIC method is its ability to discriminate between SNPs, which are not always present in the vicinity of the site. In particular, by combining PICNIC with a truncated 15-nt spacer containing crRNA we have demonstrated success in differentiating the drug-resistant K103N mutation in HIV from its wild-type counterpart, a task that is difficult to achieve with the conventional CRISPR-Cas12a due to the lack of a canonical PAM sequence. Furthermore, we also applied PICNIC for the detection and genotyping of a wide range of HCV sub-variants, particularly HCV-1a and HCV-1b, at a PAM-less site in HCV-infected human serum samples. Genotyping HCV is crucial for proper clinical management of the HCV positive individuals, however, currently it several days to get the results. Our PICNIC method demonstrated accurate detection of variants in a point-of-care setting.

The potential impact of PICNIC is not limited to infectious disease diagnostics. It can also be beneficial for other applications where the detection of genetic variants is critical, such as in cancer diagnostics, personalized medicine, as well as the detection of antimicrobial resistance in bacteria. In these fields, the ability to detect specific mutations can guide treatment decisions and allow for more effective, patient-specific interventions.

The results obtained in this study are promising and lay the groundwork for the further development and optimization of the PICNIC method. It would be interesting to explore its application in point-of-care diagnostics by coupling it with common pre-amplification methods used in the field such as RT-LAMP or reverse transcription recombinase polymerase amplification (RT-RPA) as has been previously reported by several groups, including ours. We believe our PICNIC approach can aid in the development of remarkable point-of-care tools for a wide range of applications. Although PICNIC presents a breakthrough PAM-less approach for diagnostics, other developments remain crucial for achieving unrestricted PAM requirements, particularly for genome editing in cells.

In conclusion, the PICNIC method developed here represents a significant advancement in the field of CRISPR diagnostics. By eliminating the PAM requirement and demonstrating versatility across different Cas enzymes, PICNIC expands the capabilities of CRISPR-based diagnostics and has the potential to make a substantial impact on public health.

## Methods

### Plasmid construction

Plasmids encoding Cas12a enzymes as well as the BrCas12b variants were constructed following the protocol outlined in our earlier publications^46,55,56^. A plasmid containing the *E. coli* codon-optimized Cas12i1 gene was obtained from Addgene, a gift from Arbor Biotechnologies (Plasmid #120882).

### Protein expression and purification

Rosetta bacterial colonies containing a plasmid for protein expression were initially cultured on an agar surface at 37°C overnight. Select colonies from this culture were then incubated in 10 mL of LB medium (from Fisher Scientific, #BP9723-500) for 12 hours. The culture was subsequently scaled to 1.5 liters using TB medium and continued to grow until the optical density (OD) ranged between 0.6 and 0.8. Prior to adding isopropyl β-D-1- thiogalactopyranoside (IPTG) to a final concentration of 0.5 mM, the culture was cooled on ice for about 45 to 60 minutes, followed by an additional growth period of 14 to 18 hours at 16°C.

Post-growth, the cells were collected by centrifugation at 10,000 x g for 5 minutes and resuspended in a lysis buffer containing 500 mM NaCl, 50 mM Tris-HCl (pH 7.5), 20 mM imidazole, 0.5 mM TCEP, 1 mM PMSF, 0.25 mg/mL lysozyme, and DNase I. The resuspended cells underwent sonication, followed by centrifugation at 39,800 xg for 30 minutes. The supernatant was then filtered through a 0.22 µm filter (Cytiva, #9913-2504).

For purification, the filtrate was passed through a 5 ml Histrap FF column (Cytiva, #17525501, pre-treated with Co^2+^) connected to a BioLogic DuoFlow™ FPLC system (Bio-rad), initially prepared with Wash Buffer A (500 mM NaCl, 50 mM Tris-HCl, pH 7.5, 20 mM Imidazole, 0.5 mM TCEP). Protein elution was performed using Elution Buffer B (500 mM NaCl, 50 mM

Tris-HCl, pH 7.5, 250 mM imidazole, 0.5 mM TCEP), and the eluted fractions were combined and placed in a dialysis bag (10–14 kDa MWCO) with TEV protease added (the plasmid for which was a gift from David Waugh, Addgene #8827, purified in-house). This was dialyzed in Dialysis Buffer (500 mM NaCl, 50 mM HEPES, pH 7, 5 mM MgCl2, 2 mM DTT) at 4°C overnight.

After dialysis, the protein mixture was concentrated to about 10 mL using a 30 kDa MWCO Vivaspin® 20 concentrator and balanced with 10 mL of Wash Buffer C (150 mM NaCl, 50 mM HEPES, pH 7, 0.5 mM TCEP). This was then applied to a pre-equilibrated 1 mL Hitrap Heparin HP column, part of the BioLogic DuoFlow™ FPLC system. The protein was eluted by running a gradient flow exchanging Wash Buffer C with Elution Buffer D (2000 mM NaCl, 50 mM HEPES, pH 7, 0.5 mM TCEP). Depending on purity, further purification might be required via size-exclusion chromatography using a HiLoad® 16/600 Superdex® column (Cytiva, #28989335). The purest fractions were combined, concentrated using a 30 kDa MWCO Vivaspin® 20 concentrator, and quickly frozen in liquid nitrogen before being stored at -80°C.

### Isoelectric point determination

The pI of different Cas12a, Cas12b, and Cas12i orthologs were predicted using the ProtParam tool under the Expasy suite (https://www.expasy.org/)^57^. The predicted pI of Cas12 proteins is listed in Table 2.

### Target DNA, RNA, and guide preparation

All DNA and RNA oligonucleotides, excluding the PAM library constructs, were acquired from Integrated DNA Technologies (IDT). Single-stranded oligos were diluted in 1xTE buffer (10 mM Tris, 0.1 mM EDTA, pH 7.5). Complementary oligos for synthesizing double-stranded DNA (dsDNA) were diluted in nuclease-free duplex buffer (30 mM HEPES, pH 7.5; 100 mM potassium acetate) and combined at a 1:5 molar ratio of target to non-target strand. The mixture was then denatured at 95°C for 4 minutes, followed by gradual cooling at a rate of 0.1 °C/s until reaching 25°C.

### PAM library construction

Double-stranded DNA fragments encoding different PAM sequences upstream of a conserved target sequence were ordered from Twist Biosciences to create a PAM library. The dsDNA fragments were diluted in UltraPure™ DNase/RNase-Free Distilled Water (Invitrogen #10977015) and stored at -20°C.

### Effect of temperature on dsDNA stability

A double-stranded DNA target containing a FAM label on one strand and a quencher on the complementary strand was dissolved to a final concentration of 500 nM in either nuclease-free water or PICNIC buffer. The resulting solutions were heated at different temperatures ranging from room temperature rt to 100°C for 10 min and then slowly cooled down to rt over 15 min. Fluorescence in each sample was visualized using an Analytik Jena UVP Gelstudio system under blue light (wavelength range 460–470 nm) with an ethidium bromide filter. The end-point fluorescence in each sample was quantified using a BioTek Synergy fluorescence plate reader. Fluorescence intensity measurements were recorded at excitation/emission wavelengths of 483/20 nm and 530/20 nm.

### Effect of pH on dsDNA stability

A double-stranded DNA target containing a FAM label on one strand and a quencher on the complementary strand was dissolved to a final concentration of 500 nM in nuclease-free water at different pH. The resulting solutions were heated at 85°C for 10 min and then slowly cooled down to rt over 15 min. Fluorescence visualization and quantification were performed as indicated in the ‘Effect of temperature on dsDNA stability’ section.

### Preparation of PICNIC buffer

PICNIC buffer consists of nuclease-free water substituted with 0.5% Tween-20, pH 12.

### CRISPR-Cas12a fluorescence-based trans-cleavage assay

For Cas12a and Cas12i reactions, the WT fluorescence-based detection assays without PICNIC were performed utilizing low-volume, flat-bottom, black, 384-well plates (FLUOTRAC™ 200, Greiner One). The crRNA-Cas12 conjugates were assembled in NEB 2.1 buffer and nuclease-free water and incubated for 10 minutes at room temperature. Subsequently, the assembled crRNA-Cas12 mixes were added to 500 nM FQ reporter and the appropriate concentration of the target activator in a 40 μL reaction volume. The 384-well plate containing the reaction mixture was incubated at 37°C for 30 min to 1 hour using a BioTek Synergy fluorescence plate reader. Fluorescence intensity measurements of the FAM reporter were recorded at excitation/emission wavelengths of 483/20 nm and 530/20 nm at intervals of 2.5 minutes.

For Cas12b reactions, the guide RNA-Cas12b conjugates were assembled in GeneMate 0.2 ml clear PCR strips (VWR Cat# 490003-706) at room temperature and incubated at 60°C for 10 min. Following this, 500 nM of FQ reporter and the appropriate concentration of target activator were added to the reaction to a final volume of 40 μL. The reaction was then incubated at 60°C for 30 min to 1 hour in a Bio-rad CFX96 Real-Time PCR system with a C1000 Thermal Cycler module. Unless otherwise specified, the assays were conducted with final concentrations of 60 nM Cas12a, 120 nM crRNA, and 10 nM of the target activator.

### PICNIC detection assay

The PICNIC detection assay is performed by subjecting the dsDNA target to denaturation at 85°C for 10 min in a 20 μL reaction consisting of 4 μL target, 4 μL PICNIC buffer, and 12 μL nuclease-free water. After the 10-min heating step, the 20 μL solution is added to 20 μL of CRISPR-mix consisting of 60 nM Cas12 enzyme, 120 nM crRNA, 500 nM FQ-reporter, and 1x NEB2.1 and incubated at 37°C (Cas12a and Cas12i) or 60°C (Cas12b) for 30 min to 1 hour. Fluorescence intensity measurements of the FAM reporter were recorded at excitation/emission wavelengths of 483/20 nm and 530/20 nm at intervals of 2.5 minutes.

### Patient samples extraction and processing

The collection and processing of patient samples were approved by the University of Florida Institutional Review Board (IRB202200294). For clinical validation, a total of 88 human serum samples were obtained from patients with Hepatitis C collected by HCV-TARGET. Viral RNA extraction from the serum samples was performed using the Quick-DNA/RNA Viral MagBead Kit R2140 (Zymo Research). The selection of patient samples for analysis was randomized, and the samples were blinded for one-pot testing.

The Quick-DNA/RNA Viral MagBead extraction followed the manufacturer’s protocol. Briefly, 10 µL of Proteinase K (20 mg/mL) was combined with 200 µL of serum samples in 1.5 mL centrifuge tubes and incubated at rt for 15 minutes. Subsequently, DNA/RNA ShieldTM (2x concentrate) was added to the serum sample containing Protease K at a 1:1 ratio. The combined mixture was supplemented with 800 µL of Viral DNA/RNA Buffer, followed by the addition of 20 µL of Magbinding Beads^TM^. The mixture was vortexed for 10 minutes, and the centrifuge tubes were placed in a magnetic stand to pellet the beads. The supernatant was discarded, and the beads were washed sequentially with 250 µL of MagBead DNA/RNA Wash 1, 250 µL of MagBead DNA/RNA Wash 2, and two rounds of 250 µL of 100% ethanol. After air-drying the tubes containing the beads for 10 minutes, DNA/RNA was eluted using 30 µL of DNase/RNase-Free water and subsequently subjected to a BrCas12b detection reaction.

### Genotyping of HCV-1a and HCV-1b RNA

The extracted genomic RNA from human serum patient samples were subjected to reverse transcription and amplification using Luna^®^ Probe One-Step RT-qPCR 4X Mix with UDG (NEB #M3019S) using manufacturer’s protocol and with primers targeting HCV-ns5B as described in Nakatani et.al.^54^. The amplified cDNA was subjected to detection using the PICNIC protocol and crRNA designed for HCV-1a and HCV-1b in a blind test.

## Data Availability

All the data supporting the findings of this study are available within the Article and Supplementary Files. Additional data can be obtained from the corresponding author, P.K.J., upon reasonable request.

## Acknowledgments

We thank the HCV-TARGET consortium, particularly Dr. David Nelson and Lauren Morelli, for expert guidance and clinical samples. We are grateful to the members of the Jain and Wang lab for their helpful discussions and to the University of Florida (UF) Health Cancer Center for their support. This work was financially supported by funds from the UF, UF Herbert Wertheim College of Engineering, Shah Foundation Endowment Funds, NIH-NIAID R21AI156321, NIH-NIAID R21AI168795, and NIH-NIGMS R35GM147788. The funding sources did not have a role in the design of the study, the collection, analysis, or interpretation of data, nor in writing the manuscript. All schematics were created with Biorender.com.

## References

1. Baker, R. E. et al. Infectious disease in an era of global change. Nat. Rev. Microbiol. 20, 193–205 (2022).

2. Fauci, A. S. Infectious Diseases: Considerations for the 21st Century. Clin. Infect. Dis. 32, 675–685 (2001).

3. Kaminski, M. M., Abudayyeh, O. O., Gootenberg, J. S., Zhang, F. & Collins, J. J. CRISPR-based diagnostics. Nat. Biomed. Eng. 5, 643–656 (2021).

4. Kulkarni, A., Tanga, S., Karmakar, A., Hota, A. & Maji, B. CRISPR-Based Precision Molecular Diagnostics for Disease Detection and Surveillance. ACS Appl. Bio Mater. 6, 3927–3945 (2023).

5. Kellner, M. J., Koob, J. G., Gootenberg, J. S., Abudayyeh, O. O. & Zhang, F. SHERLOCK: nucleic acid detection with CRISPR nucleases. Nat. Protoc. 14, 2986–3012 (2019).

6. Broughton, J. P. et al. CRISPR–Cas12-based detection of SARS-CoV-2. Nat. Biotechnol. 38, 870–874 (2020).

7. Long T. Nguyen, Brianna M. Smith, & Piyush K. Jain. Enhancement of trans-cleavage activity of Cas12a with engineered crRNA enables amplified nucleic acid detection. *bioRxiv* 2020.04.13.036079 (2020) doi:10.1101/2020.04.13.036079.

8. Nguyen, L. T. et al. A thermostable Cas12b from Brevibacillus leverages one-pot discrimination of SARS-CoV-2 variants of concern. eBioMedicine 77, (2022).

9. Jinek, M. et al. A Programmable Dual-RNA–Guided DNA Endonuclease in Adaptive Bacterial Immunity. Science 337, 816–821 (2012).

10. Pinilla-Redondo, R. et al. Type IV CRISPR–Cas systems are highly diverse and involved in competition between plasmids. Nucleic Acids Res. 48, 2000–2012 (2020).

11. Makarova, K. S. et al. Evolutionary classification of CRISPR–Cas systems: a burst of class 2 and derived variants. Nat. Rev. Microbiol. 18, 67–83 (2020).

12. Chen, J. S. et al. CRISPR-Cas12a target binding unleashes indiscriminate single-stranded DNase activity. Science 360, 436–439 (2018).

13. Hajizadeh Dastjerdi, A., Newman, A. & Burgio, G. The Expanding Class 2 CRISPR Toolbox: Diversity, Applicability, and Targeting Drawbacks. BioDrugs 33, 503–513 (2019).

14. Gleditzsch, D. et al. PAM identification by CRISPR-Cas effector complexes: diversified mechanisms and structures. RNA Biol. 16, 504–517 (2019).

15. Leenay, R. T. & Beisel, C. L. Deciphering, Communicating, and Engineering the CRISPR PAM. J. Mol. Biol. 429, 177–191 (2017).

16. Tóth, E. et al. Improved LbCas12a variants with altered PAM specificities further broaden the genome-targeting range of Cas12a nucleases. Nucleic Acids Res. 48, 3722–3733 (2020).

17. Creutzburg, S. C. A. et al. Good guide, bad guide: spacer sequence-dependent cleavage efficiency of Cas12a. Nucleic Acids Res. 48, 3228–3243 (2020).

18. Collias, D. & Beisel, C. L. CRISPR technologies and the search for the PAM-free nuclease. Nat. Commun. 12, 555 (2021).

19. Ghouneimy, A., Mahas, A., Marsic, T., Aman, R. & Mahfouz, M. CRISPR-Based Diagnostics: Challenges and Potential Solutions toward Point-of-Care Applications. ACS Synth. Biol. 12, 1–16 (2023).

20. Duzkale, H. et al. A systematic approach to assessing the clinical significance of genetic variants. Clin. Genet. 84, 453–463 (2013).

21. Hart, J. R. et al. The butterfly effect in cancer: A single base mutation can remodel the cell. Proc. Natl. Acad. Sci. 112, 1131–1136 (2015).

22. Shigeto, H. et al. Analysis of Single Nucleotide-Mutated Single-Cancer Cells Using the Combined Technologies of Single-Cell Microarray Chips and Peptide Nucleic Acid-DNA Probes. Micromachines 11, (2020).

23. Zhao, H. et al. First Detection in West Africa of a Mutation That May Contribute to Artemisinin Resistance Plasmodium falciparum. Front. Genet. 12, (2021).

24. Shi, K. et al. PAM-free cascaded strand displacement coupled with CRISPR-Cas12a for amplified electrochemical detection of SARS-CoV-2 RNA. Anal. Biochem. 664, 115046 (2023).

25. Jiang, W., Aman, R., Ali, Z., Rao, G. S. & Mahfouz, M. PNA-Pdx: Versatile Peptide Nucleic Acid-Based Detection of Nucleic Acids and SNPs. Anal. Chem. 95, 14209–14218 (2023).

26. Wu, Y. et al. A PAM-free CRISPR/Cas12a ultra-specific activation mode based on toehold-mediated strand displacement and branch migration. Nucleic Acids Res. 50, 11727– 11737 (2022).

27. Zhu, Z. et al. PAM-free loop-mediated isothermal amplification coupled with CRISPR/Cas12a cleavage (Cas-PfLAMP) for rapid detection of rice pathogens. Biosens. Bioelectron. 204, 114076 (2022).

28. Zhou, S. et al. Endonuclease-Assisted PAM-free Recombinase Polymerase Amplification Coupling with CRISPR/Cas12a (E-PfRPA/Cas) for Sensitive Detection of DNA Methylation. ACS Sens. 7, 3032–3040 (2022).

29. Hu, Y. et al. Development of an inducible Cas9 nickase and PAM-free Cas12a platform for bacterial diagnostics. Talanta 265, 124931 (2023).

30. Nishimasu, H. et al. Structural Basis for the Altered PAM Recognition by Engineered CRISPR-Cpf1. Mol. Cell 67, 139–147.e2 (2017).

31. Yamano, T. et al. Structural Basis for the Canonical and Non-canonical PAM Recognition by CRISPR-Cpf1. Mol. Cell 67, (2017).

32. Jacobsen, T., Liao, C. & Beisel, C. L. The Acidaminococcus sp. Cas12a nuclease recognizes GTTV and GCTV as non-canonical PAMs. FEMS Microbiol. Lett. 366, fnz085 (2019).

33. Jacobsen, T. et al. Characterization of Cas12a nucleases reveals diverse PAM profiles between closely-related orthologs. Nucleic Acids Res. 48, 5624–5638 (2020).

34. Zetsche, B., Abudayyeh, O. O., Gootenberg, J. S., Scott, D. A. & Zhang, F. A Survey of Genome Editing Activity for 16 Cas12a Orthologs. Keio J. Med. 69, 59–65 (2020).

35. Vasilev, R. et al. Targeted Modification of Mammalian DNA by a Novel Type V Cas12a Endonuclease from Ruminococcus bromii. Int. J. Mol. Sci. 23, (2022).

36. Zhang, J. et al. Efficient Multiplex Genome Editing in Streptomyces via Engineered CRISPR-Cas12a Systems. Front. Bioeng. Biotechnol. 8, (2020).

37. Kleinstiver, B. P. et al. Engineered CRISPR–Cas12a variants with increased activities and improved targeting ranges for gene, epigenetic and base editing. Nat. Biotechnol. 37, 276–282 (2019).

38. Zhang, Y. et al. Expanding the scope of plant genome engineering with Cas12a orthologs and highly multiplexable editing systems. Nat. Commun. 12, 1944 (2021).

39. Lin, Q. et al. Genome editing in plants with MAD7 nuclease. J. Genet. Genomics 48, 444–451 (2021).

40. 40. Chen, Y., et al. Synergistic engineering of CRISPR-Cas nucleases enables robust mammalian genome editing. The Innovation 3, (2022).

41. Zetsche, B. et al. Cpf1 Is a Single RNA-Guided Endonuclease of a Class 2 CRISPR-Cas System. Cell 163, 759–771 (2015).

42. Paul, B. & Montoya, G. CRISPR-Cas12a: Functional overview and applications. Biomed. J. 43, 8–17 (2020).

43. Shmakov, S. et al. Diversity and evolution of class 2 CRISPR–Cas systems. Nat. Rev. Microbiol. 15, 169–182 (2017).

44. Shmakov, S. et al. Discovery and Functional Characterization of Diverse Class 2 CRISPR-Cas Systems. Mol. Cell 60, 385–397 (2015).

45. Zhang, H., Li, Z., Xiao, R. & Chang, L. Mechanisms for target recognition and cleavage by the Cas12i RNA-guided endonuclease. Nat. Struct. Mol. Biol. 27, 1069–1076 (2020).

46. Nguyen, L. T. et al. Engineering highly thermostable Cas12b via de novo structural analyses for one-pot detection of nucleic acids. Cell Rep. Med. 4, (2023).

47. Yan, W. X. et al. Functionally diverse type V CRISPR-Cas systems. Science 363, 88– 91 (2019).

48. Joung, J. et al. Detection of SARS-CoV-2 with SHERLOCK one-pot testing. N. Engl. J. Med. 383, 1492–1494 (2020).

49. Akinsete, O. et al. K103N Mutation in Antiretroviral Therapy—Naive African Patients Infected with HIV Type 1. Clin. Infect. Dis. 39, 575–578 (2004).

50. Wang, Y. et al. The development of drug resistance mutations K103N Y181C and G190A in long term Nevirapine-containing antiviral therapy. AIDS Res. Ther. 11, 36 (2014).

51. Stumpf, M. P. H. & Pybus, O. G. Genetic diversity and models of viral evolution for the hepatitis C virus. FEMS Microbiol. Lett. 214, 143–152 (2002).

52. Martinez, M. A. & Franco, S. Therapy Implications of Hepatitis C Virus Genetic Diversity. Viruses 13, (2021).

53. Pellicelli, A. M. et al. HCV genotype 1a shows a better virological response to antiviral therapy than HCV genotype 1b. BMC Gastroenterol. 12, 162 (2012).

54. Nakatani, S. M. et al. Development of Hepatitis C Virus Genotyping by Real-Time PCR Based on the NS5B Region. PLOS ONE 5, e10150 (2010).

55. Rananaware, S. R. et al. Programmable RNA detection with CRISPR-Cas12a. Nat. Commun. 14, 5409 (2023).

56. Nguyen, L. T. et al. Harnessing noncanonical crRNAs to improve functionality of Cas12a orthologs. Cell Rep. 43, (2024).

57. Gasteiger, E. et al. Protein Identification and Analysis Tools on the Expasy Server. In The Proteomics Protocols Handbook (ed. Walker, J. M.) 571–607 (Humana Press, 2005).

58. Jumper, J. et al. Highly accurate protein structure prediction with AlphaFold. Nature 596, 583–589 (2021).

59. Zhang, B. et al. Mechanistic insights into the R-loop formation and cleavage in CRISPR-Cas12i1. Nat. Commun. 12, 3476 (2021).

