## Supplementary File for "PAM-free diagnostics with diverse type V CRISPR-Cas systems"

---

### Supplementary Information

---

**Table 1:** List of protein sequences used in this study:

| Sr. No. | Name | Sequence |
|---|---|---|
| 1 | LbCas12a | MSKLEKFTNCYSLSKTLRFKAIPVGKTQENIDNKRLLVEDEKRAEDYKGVKKLL<br>DRYYLSFINDVLHSIKLKNLNNYISLFRKKTRTEKENKELENLEINLRKEIAKAFK<br>GNEGYKSLFKKDIIETILPEFLDDKDEIALVNSFNGFTTAFTGFFDNRENMFSEEA<br>KSTSIAFRCINENLTRYISNMDIFEKVDAIFDKHEVQEIKEKILNSDYDVEDFFEGE<br>FFNFVLTQEGIDVYNAIIGGFVTESGEKIKGLNEYINLYNQKTKQKLPKFKPLYKQ<br>VLSDRESLSFYGEGYTSDEEVLEVFRNTLNKNSEIFSSIKKLEKLFKNFDEYSSAGI<br>FVKNGPAISTISKDIFGEWNVIRDKWNAEYDDIHLKKKAVVTEKYEDDRRKSFK<br>KIGSFSLEQLQEYADADLSVVEKLKEIIIQKVDEIYKVYGSSEKLFDADFVLEKSL<br>KKNDAVVAIMKDLLDSVKSFENYIKAFFGEGKETNRDESFYGDFVLAYDILLKV<br>DHIYDAIRNYVTQKPYSKDKFKLYFQNPQFMGGWDKDKETDYRATILRYGSKY<br>YLAIMDKKYAKCLQKIDKDDVNGNYEKINYKLLPGPNKMLPKVFFSKKWMAY<br>YNPSEDIQKIYKNGTFKKGDMFNLNDCHKLIDFFKDSISRYPKWSNAYDFNFSET<br>EKYKDIAGFYREVEEQGYKVSFESASKKEVDKLVEEGKLYMFQIYNKDFSDKSH<br>GTPNLHTMYFKLLFDENNHGQIRLSGGAELFMRRASLKKEELVVHPANSPIANK<br>NPDNPKKTTTLSYDVYKDKRFSEDQYELHIPIAINKCPKNIFKINTEVRVLLKHDD<br>NPYVIGIDRGERNLLYIVVVDGKGNIVEQYSLNEIINNFNGIRIKTDYHSLLDKKE<br>KERFEARQNWTSIENIKELKAGYISQVVHKICELVEKYDAVIALEDLNSGFKNSR<br>VKVEKQVYQKFEKMLIDKLNYMVDKKSNPCATGGALKGYQITNKFESFKSMST<br>QNGFIFYIPAWLTSKIDPSTGFVNLLKTKYTSIADSKKFISSFDRIMYVPEEDLFEF<br>ALDYKNFSRTDADYIKKWKLYSYGNRIRIFRNPKKNNVFDWEEVCLTSAYKELF<br>NKYGINYQQGDIRALLCEQSDKAFYSSFMALMSLMLQMRNSITGRTDVDFLISP<br>VKNSDGIFYDSRNYEAQENAILPKNADANGAYNIARVLWAIGQFKKAEDEKLD<br>KVKIAISNKEWLEYAQTSVKH |
| 2 | AsCas12a | MTQFEGFTNLYQVSKTLRFELIPQGKTLKHIQEQGFIEEDKARNDHYKELKPIIDR<br>IYKTYADQCLQLVQLDWENLSAAIDSYRKEKTEETRNALIEEQATYRNAIHDYFI<br>GRTDNLTDAINKRHAEIYKGLFKAELFNGKVLKQLGTVTTTEHENALLRSFDKF<br>TTYFSGFYENRKNVFSAEDISTAIPHRIVQDNFPKFKENCHIFTRLITAVPSLREHF<br>ENVKKAIGIFVSTSIEEVFSFPFYNQLLTQTQIDLYNQLLGGISREAGTEKIKGLNE<br>VLNLAIQKNDETAHIIASLPHRFIPLFKQILSDRNTLSFILEEFKSDEEVIQSFCKYK<br>TLLRNENVLETAEALFNELNSIDLTHIFISHKKLETISSALCDHWDTLRNALYERRI<br>SELTGKITKSAKEKVQRSLKHEDINLQEIISAAGKELSEAFKQKTSEILSHAHAAL<br>DQPLPTTLKKQEEKEILKSQLDSLLGLYHLLDWFAVDESNEVDPEFSARLTGIKL<br>EMEPSLSFYNKARNYATKKPYSVEKFKLNFQMPTLASGWDVNKEKNNGAILFV<br>KNGLYYLGIMPKQKGRYKALSFEPTEKTSEGFDKMYYDYFPDAAKMIPKCSTQL<br>KAVTAHFQTHTTPILLSNNFIEPLEITKEIYDLNNPEKEPKKFQTAYAKKTGDQKG<br>YREALCKWIDFTRDFLSKYTKTTSIDLSSLRPSSQYKDLGEYYAELNPLLYHISFQ<br>RIAEKEIMDAVETGKLYLFQIYNKDFAKGHHGKPNLHTLYWTGLFSPENLAKTSI<br>KLNGQAELFYRPKSRMKRMAHRLGEKMLNKKLKDQKTPIPDTLYQELYDYVN<br>HRLSHDLSDEARALLPNVITKEVSHEIIKDRRFTSDKFFFHVPITLNYQAANSPSKF<br>NQRVNAYLKEHPETPIIGIDRGERNLIYITVIDSTGKILEQRSLNTIQQFDYQKKLD<br>NREKERVAARQAWSVVGTIKDLKQGYLSQVIHEIVDLMIHYQAVVVLENLNFGF<br>KSKRTGIAEKAVYQQFEKMLIDKLNCLVLKDYPAEKVGGVLNPYQLTDQFTSFA<br>KMGTQSGFLFYVPAPYTSKIDPLTGFVDPFVWKTIKNHESRKHFLEGFDFLHYDV<br>KTGDFILHFKMNRNLSFQRGLPGFMPAWDIVFEKNETQFDAKGTPFIAGKRIVPV<br>IENHRFTGRYRDLYPANELIALLEEKGIVFRDGSNILPKLLENDDSHAIDTMVALI<br>RSVLQMRNSNAATGEDYINSPVRDLNGVCFDSRFQNPEWPMDADANGAYHIAL<br>KGQLLLNHLKESKDLKLQNGISNQDWLAYIQELRN |

| | | |
|---|---|---|
| 3 | ErCas12a | NNGTNNFQNFIGISSLQKTLRNALIPTETTQQFIVKNGIIKEDELRGENRQILKDIM<br>DDYYRGFISETLSSIDDIDWTSLFEKMEIQLKNGDNKDTLIKEQTEYRKAIHKKFA<br>NDDRFKNMFSAKLISDILPEFVIHNNNYSASEKEEKTQVIKLFSRFATSFKDYFKN<br>RANCFSADDISSSSCHRIVNDNAEIFFSNALVYRRIVKSLSNDDINKISGDMKDSL<br>KEMSLEEIYSYEKYGEFITQEGISFYNDICGKVNSFMNLYCQKNKENKNLYKLQK<br>LHKQILCIADTSYEVPYKFESDEEVYQSVNGFLDNISSKHIVERLRKIGDNYNGYN<br>LDKIYIVSKFYESVSQKTYRDWETINTALEIHYNNILPGNGKSKADKVKKAVKN<br>DLQKSITEINELVSNYKLCSDDNIKAETYIHEISHILNNFEAQELKYNPEIHLVESE<br>LKASELKNVLDVIMNAFHWCSVFMTEELVDKDNNFYAELEEIYDEIYPVISLYNL<br>VRNYVTQKPYSTKKIKLNFGIPTLADGWSKSKEYSNNAIILMRDNLYYLGIFNAK<br>NKPDKKIIEGNTSENKGDYKKMIYNLLPGPNKMIPKVFLSSKTGVETYKPSAYIL<br>EGYKQNKHIKSSKDFDITFCHDLIDYFKNCIAIHPEWKNFGFDFSDTSTYEDISGF<br>YREVELQGYKIDWTYISEKDIDLLQEKGQLYLFQIYNKDFSKKSTGNDNLHTMY<br>LKNLFSEENLKDIVLKLNGEAEIFFRKSSIKNPIIHKKGSILVNRTYEAEEKDQFGN<br>IQIVRKNIPENIYQELYKYFNDKSDKELSDEAAKLKNVVGHHEAATNIVKDYRYT<br>YDKYFLHMPITINFKANKTGFINDRILQYIAKEKDLHVIGIDRGERNLIYVSVIDTC<br>GNIVEQKSFNIVNGYDYQIKLKQREGARQIARKEWKEIGKIKEIKEGYLSLVIHEI<br>SKMVIKYNAIIVMEDLSYGFKKGRFKVERQVYQKFETMLINKLNYLVFKDISITE<br>NGGLLKGYQLTYIPDKLKNVGHQCGCIFYVPAAYTSKIDPTTGFVNIFKFKDLTV<br>DAKREFIKKFDSIRYDSEKNLFCFTFDYNNFITQNTVMSKSSWSVYTYGVRIKRR<br>FVNGRFSNESDTIDITKDMEKTLEMTDINWRDGHDLRQDIIDYEIVQHIFEIFRLT<br>VQMRNSLSELEDRDYDRLISPVLNENNIFYDSAKAGDALPKDADANGAYCIALK<br>GLYEIKQITENWKEDGKFSRDKLKISNKDWFDFIQNKRYL |
| 4 | BrCas12b RFND | MPVRSFKVKLVTRSGDAEHMLQLRRGLWKTHEIVNQGIAYYMNKLALMRQEP<br>YAGKSREVVRLELLHSLRAQQKRNNWTGDAGTDDEILNLSRRLYELLVPSAIGE<br>KGDAQMLSRKFLSPLVDPNSEGGKGTAKSGRKPRWMKMREEGHPDWEAEREK<br>DEAKKAADPTASILNDLEAFGLRPLFPLFTDEQKGIQWLPKQKRQFVRTWDRDM<br>FQQALERMLSWESWNRRVAEEYQKLQAQRDELYAKYLADGGAWLEALQSFEK<br>QREVELAEESFAAKSEYLITRRQIRGWKQVYEKWSQLPEHAAQEQFWQVVADV<br>QTSLPGAFGDPKVYQFLSQPEHHHIWRGYPNRLFHYSDYNGVRKKLQRARHDA<br>TFTLPDPVEHPLWIRFDARGGNIHDYEISQNGKQYQVTFSRLLWPENETWVERE<br>NVTVAIGASQQLKRQIRLDGYADKKQKVRYRDYSSGIELTGVLGGAKIQFDRRH<br>LRKASNRLADGETGPVYLNVVVDIEPFLAMRNGRLQTPIGQVLQVVTKDWPKV<br>TGYKPAELISWIQNSPLAVGTGVNTIEAGMRVMSVDLGQRSAAAVSIFEVMRQK<br>PAEQETKLFYPIAVTGLYAVHRRSLLLRLPGEKISDEIEQQRKIRAHARSLVRYQI<br>RLLADVLRLHTRGTAEQRRAKLDELLATLQTKQELDQKLWQTELEKLFDYIHEP<br>AERWQQALVAAHRTLEPVIGQAVRHWRKSLRIDRKGLAGMSMWNIEELEETRK<br>LLIAWSKHSRVPGEPNRLDKEETFAPQQLQHIQNVKDDRLKQMANLLVMTALG<br>YKYDEAEKQWKEAYPACQMILFEDLSRYRFALDRPRRENNRLMKWAHRSIPRL<br>VYLQGELFGIQVGVVYSAYTSRFHAKTGAPGIRCHALKEEDLQPNSYVVKQLIK<br>DGFIREDQTGSLKPGQIVPWSGGELFVTLADRSGSRLAVIHADINAAQNLQKRFW<br>QQNTEIFRVPCKVTTSGLIPAYDKMKKLFGKGYFAKINQTDTSEVYVWEHSAKM<br>KGKTTPADPAEEGVFDESLTDEMEELEDSQEGYKTLFRDPSGFFWSSDRWLPQK<br>EFWFWVKRRIEKKLREQLQ |
| 5 | Cas12i1 | MSNKEKNASETRKAYTTKMIPRSHDRMKLLGNFMDYLMDGTPIFFELWNQFGG<br>GIDRDIISGTANKDKISDDLLLAVNWFKVMPINSKPQGVSPSNLANLFQQYSGSEP<br>DIQAQEYFASNFDTEKHQWKDMRVEYERLLAELQLSRSDMHHDLKLMYKEKCI<br>GLSLSTAHYITSVMFGTGAKNNRQTKHQFYSKVIQLLEESTQINSVEQLASIILKA<br>GDCDSYRKLRIRCSRKGATPSILKIVQDYELGTNHDDEVNVPSLIANLKEKLGRF<br>EYECEWKCMEKIKAFLASKVGPYYLGSYSAMLENALSPIKGMTTKNCKFVLKQI<br>DAKNDIKYENEPFGKIVEGFFDSPYFESDTNVKWVLHPHHIGESNIKTLWEDLNA<br>IHSKYEEDIASLSEDKKEKRIKVYQGDVCQTINTYCEEVGKEAKTPLVQLLRYLY<br>SRKDDIAVDKIIDGITFLSKKHKVEKQKINPVIQKYPSFNFGNNSKLLGKIISPKDK<br>LKHNLKCNRNQVDNYIWIEIKVLNTKTMRWEKHHYALSSTRFLEEVYYPATSEN<br>PPDALAARFRTKTNGYEGKPALSAEQIEQIRSAPVGLRKVKKRQMRLEAARQQN<br>LLPRYTWGKDFNINICKRGNNFEVTLATKVKKKKEKNYKVVLGYDANIVRKNT<br>YAAIEAHANGDGVIDYNDLPVKPIESGFVTVESQVRDKSYDQLSYNGVKLLYCK |

| | | |
|---|---|---|
| | | PHVESRRSFLEKYRNGTMKDNRGNNIQIDFMKDFEAIADDETSLYYFNMKYCKL<br>LQSSIRNHSSQAKEYREEIFELLRDGKLSVLKLSSLSNLSFVMFKVAKSLIGTYFG<br>HLLKKPKNSKSDVKAPPITDEDKQKADPEMFALRLALEEKRLNKVKSKKEVIAN<br>KIVAKALELRDKYGPVLIKGENISDTTKKGKKSSTNSFLMDWLARGVANKVKE<br>MVMMHQGLEFVEVNPNFTSHQDPFVHKNPENTFRARYSRCTPSELTEKNRKEIL<br>SFLSDKPSKRPTNAYYNEGAMAFLATYGLKKNDVLGVSLEKFKQIMANILHQRS<br>EDQLLFPSRGGMFYLATYKLDADATSVNWNGKQFWVCNADLVAAYNVGLVDI<br>QKDFKKK |

**Table 2:** List of Cas12a orthologs and their isoelectric points (Red = high pI, Blue = low pI)

| **Sr. No.** | **Name** | **Isoelectric Point** |
|---|---|---|
| 1 | CmtCas12a | 8.72 |
| 2 | Mb2Cas12a | 8.66 |
| 3 | PdCas12a | 8.58 |
| 4 | FnCas12a | 8.57 |
| 5 | MbCas12a | 8.47 |
| 6 | Mb3Cas12a | 8.47 |
| 7 | LbCas12a | 8.38 |
| 8 | HkCas12a | 8.36 |
| 9 | Pb2Cas12a | 8.28 |
| 10 | PcCas12a | 8.2 |
| 11 | BoCas12a | 8.18 |
| 12 | AsCas12a | 8.01 |
| 13 | BfCas12 | 7.76 |
| 14 | ArCas12a | 7.72 |
| 15 | MICas12a | 7.65 |
| 16 | TsCas12a | 7.15 |
| 17 | Lb5Cas12a | 7.04 |
| 18 | ErCas12a | 6.71 |
| 19 | BsCas12a | 6.47 |
| 20 | LpCas12a | 6.38 |
| 21 | CmaCas12a | 6.09 |
| 22 | PxCas12a | 5.66 |
| 23 | PrCas12a | 5.57 |

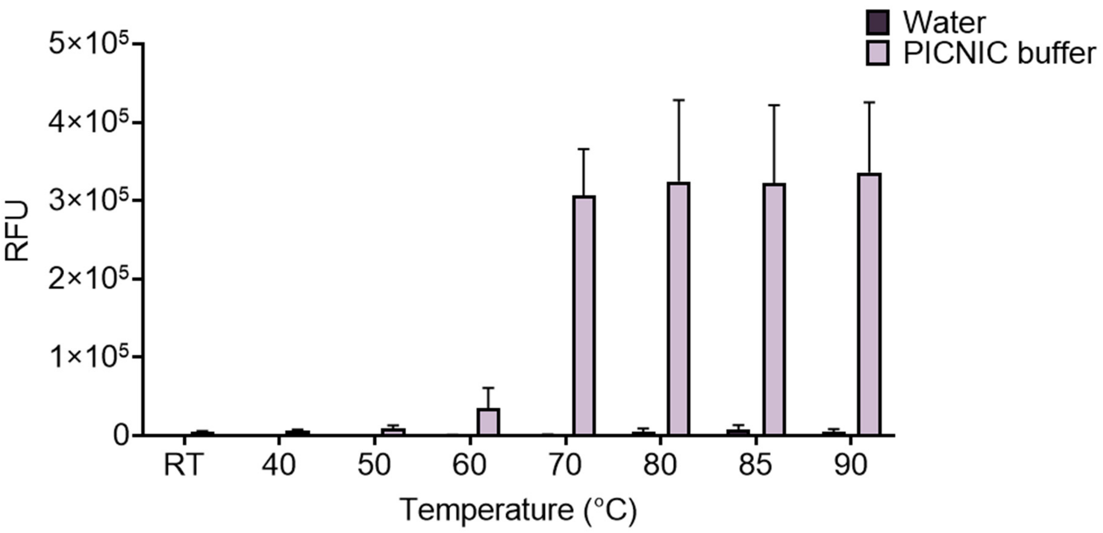


**Fig. S1:** Temperature dependency of denaturation of dsDNA substrates. Data showing the temperature dependence of trans-cleavage-mediated fluorescence activity in RFU with and without the addition of PICNIC buffer (pH=12) and temperatures ranging from room temperature (RT) to 90°C. Error bars represent Mean ± S.D. (n=3).

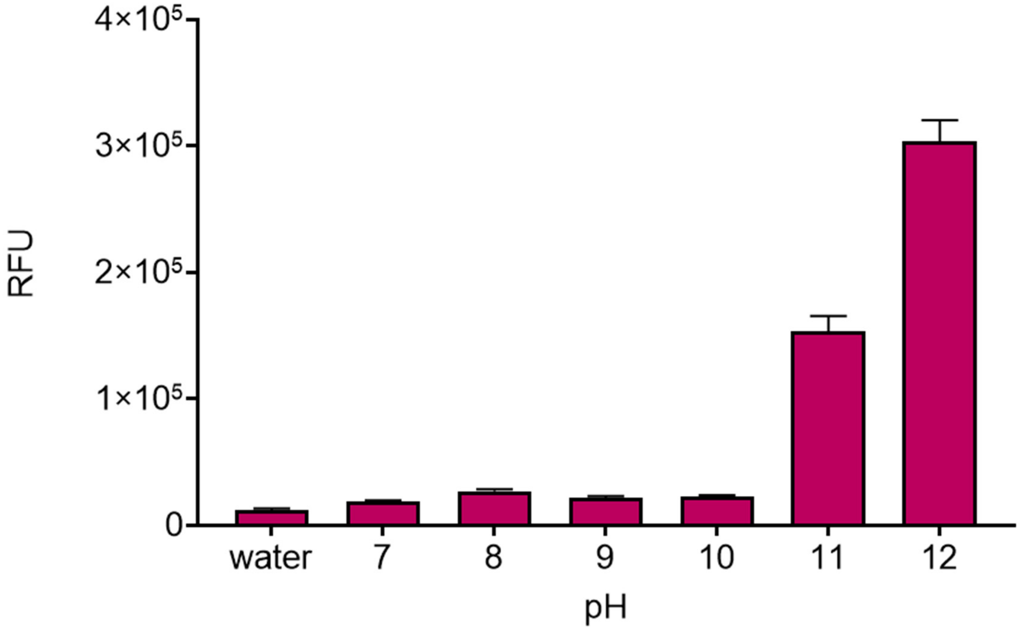


**Fig. S2:** The effect of pH on the DNA denaturation dynamics. Data showing trans cleavage dependence on pH for a PICNIC reaction. The pH ranges from neutral (7) to 12, with water used as a control. Plot represents the fluorescence intensity in RFU at each condition. Error bars represent Mean ± S.D. (n=3).

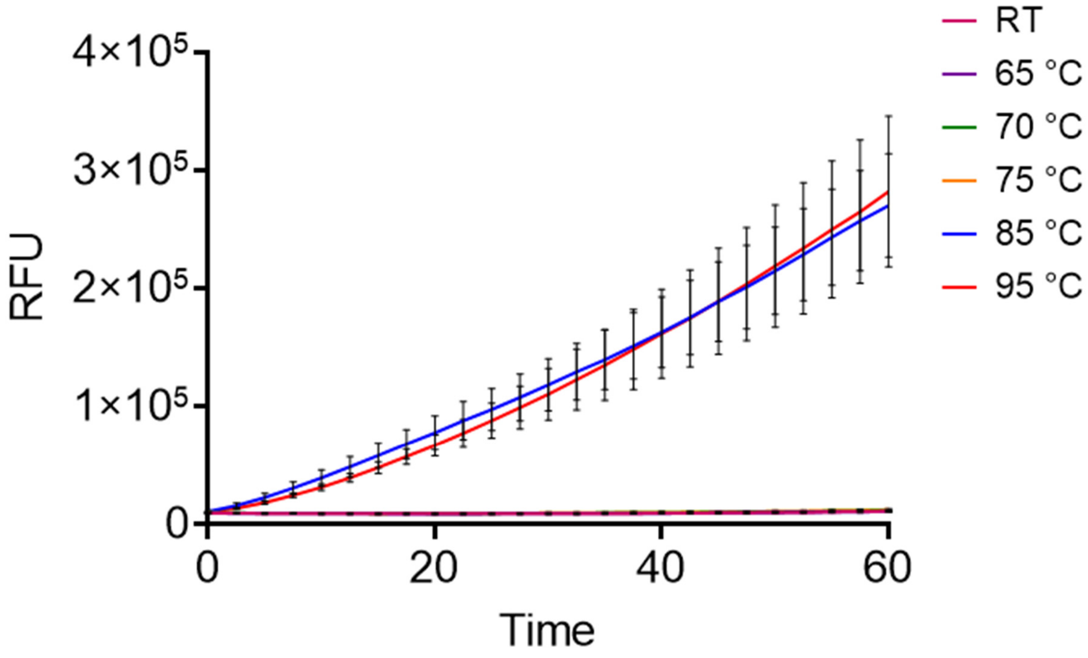


**Fig. S3:** The effect of incubation temperature on PICNIC method. Graph depicting trans-cleavage fluorescence values utilizing the PICNIC method at various temperatures, ranging from room temperature (RT) to 95°C. Error bars represent Mean ± SD (n=3) is indicated.

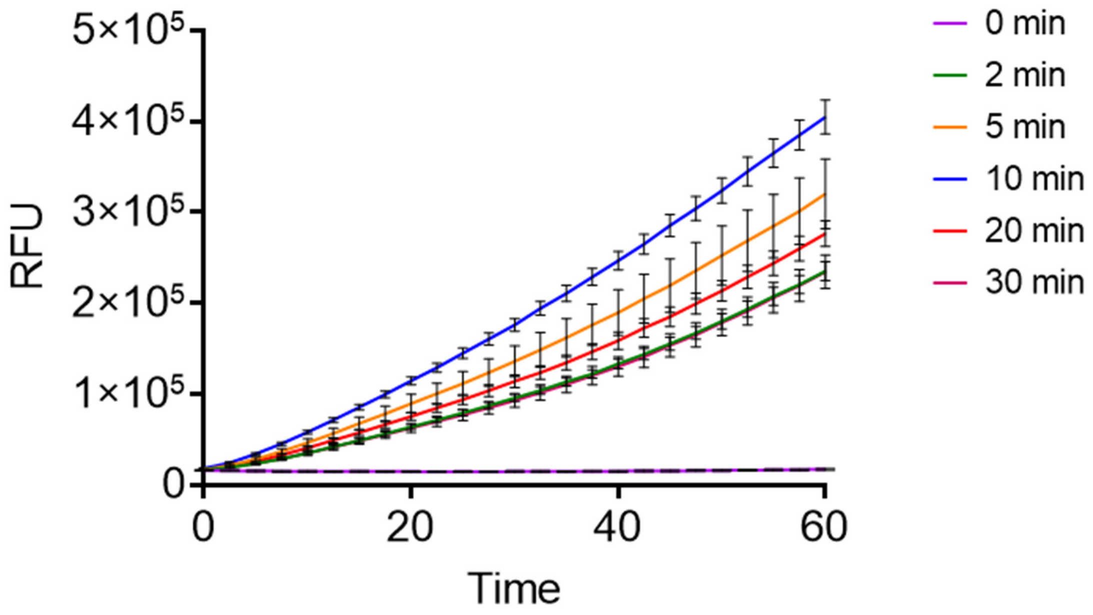


**Fig. S4:** The effect of heating time on PICNIC method**.** Graph illustrating the time dependence of target dsDNA denaturation via incubation, from 0 min to 30 min. All samples are incubated in PICNIC buffer at a pH of 12 and a temperature of 100°C and cooled for 15 minutes at room temperature. Error bars represent Mean ± SD (n = 3).

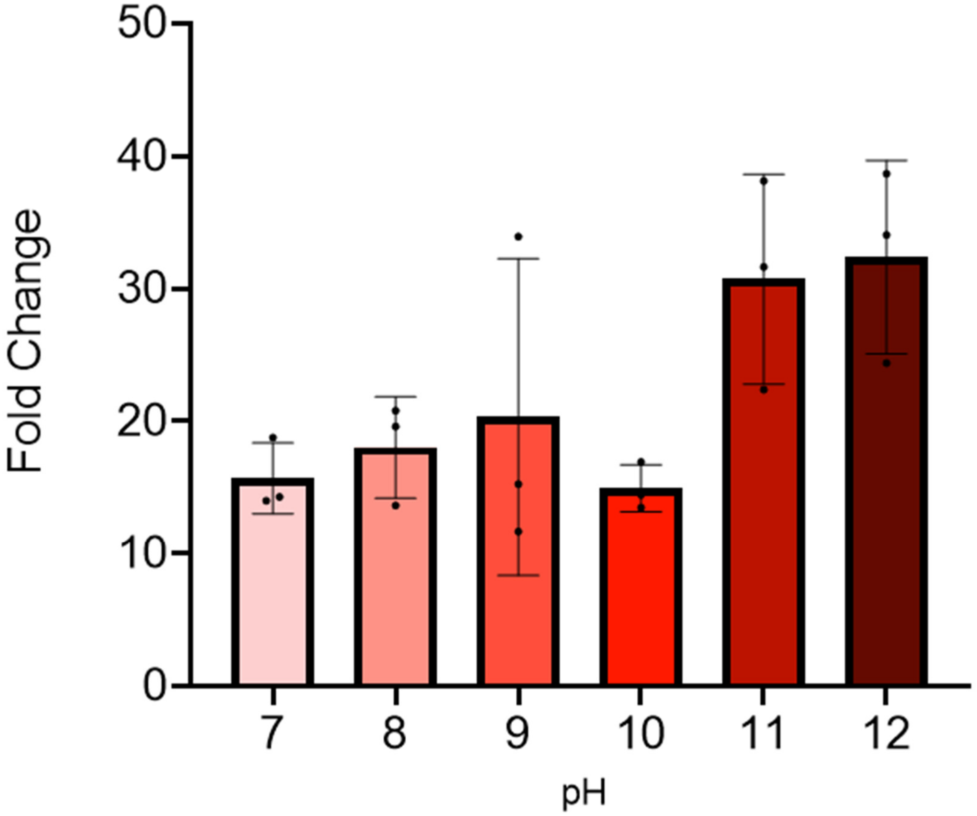


**Fig. S5:** pH dependence of trans-cleavage activity using the PICNIC method. All samples are incubated under the same conditions at 100°C for 10 minutes with differing pH. pH levels left to right are 7, 8, 9, 10, 11, and 12. Mean ± SD (n = 3) is indicated.

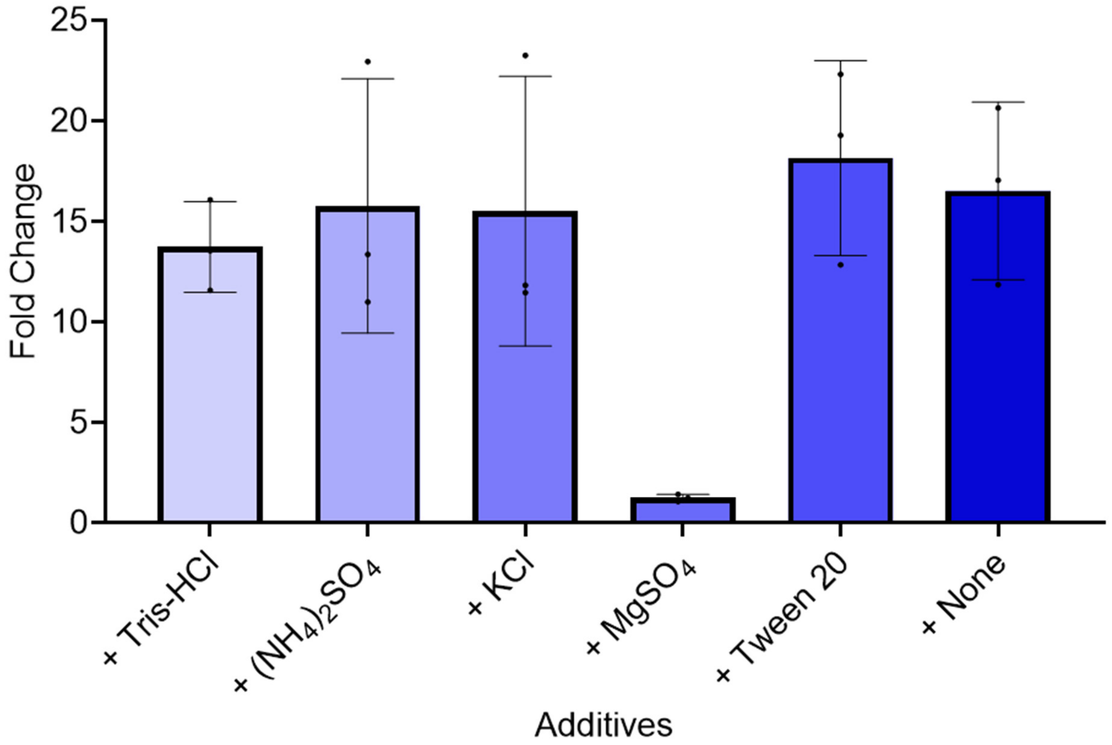


**Fig. S6:** Fold change at t=30 minutes utilizing the PICNIC method using water at a pH of 12. Various additives to this water at a pH of 12 are indicated. Mean ± SD (n = 3) is indicated.

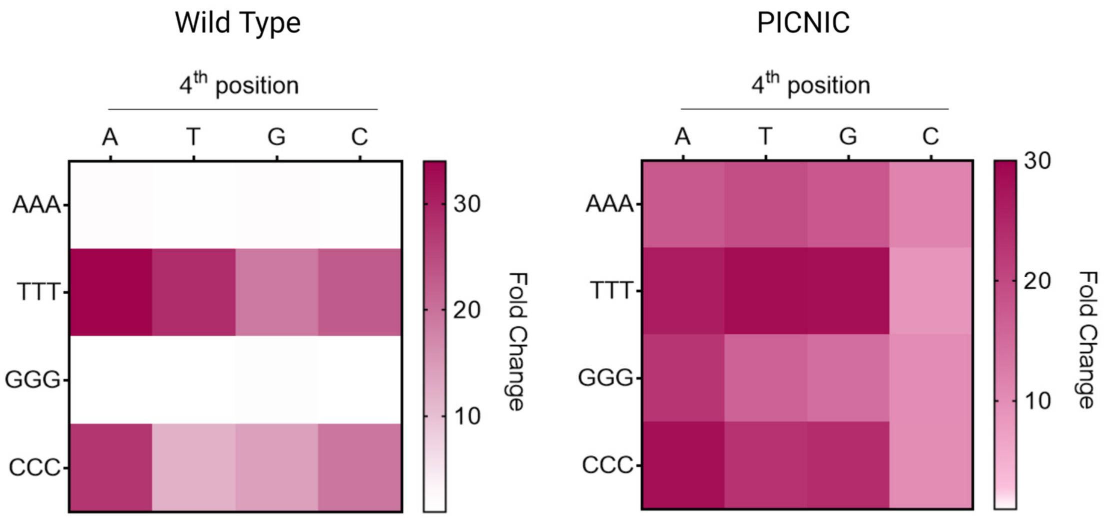


**Fig. S7:** The effect of changing the 4th nucleotide in the PAM sequence with WT-CRISPR and PICNIC method. Figure showing trans-cleavage activity of target sequences preceded by various PAM sequences under both Wild Type and PICNIC conditions. In this figure the fourth nucleotide of the 4-nucleotide PAM sequence is varied.

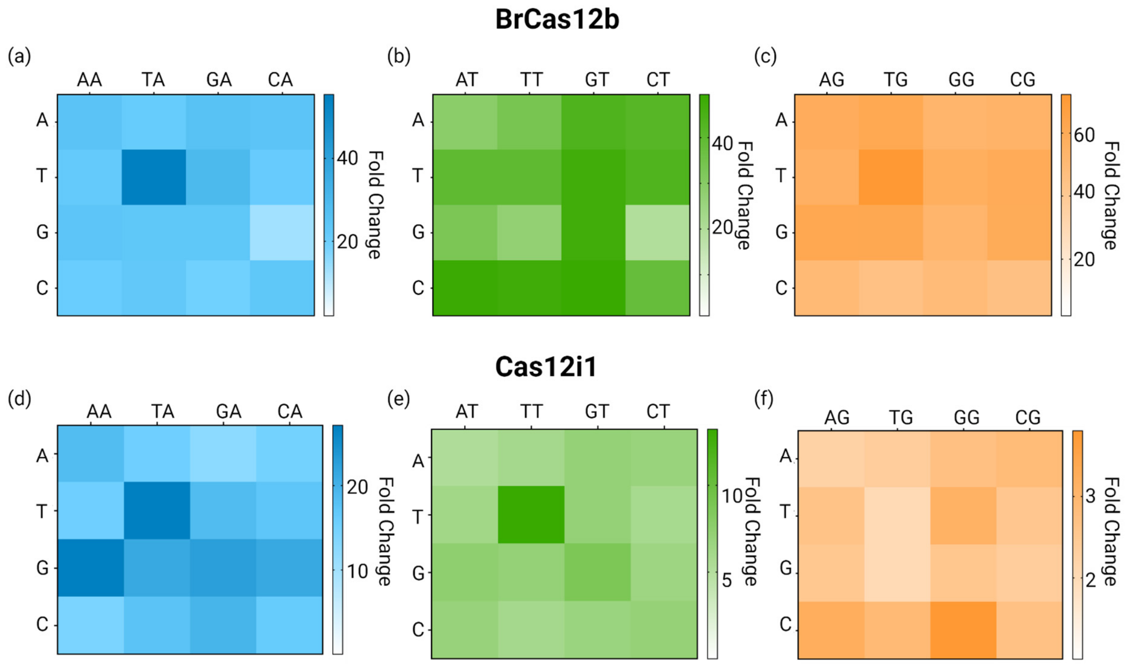


**Fig. S8:** PAM-library trans-cleavage with Cas12b and Cas12i. Figure showing trans-cleavage activity of target sequences preceded by various 'NNN' PAM sequences with PICNIC conditions using BrCas12b (top) and Cas12i1 (bottom).

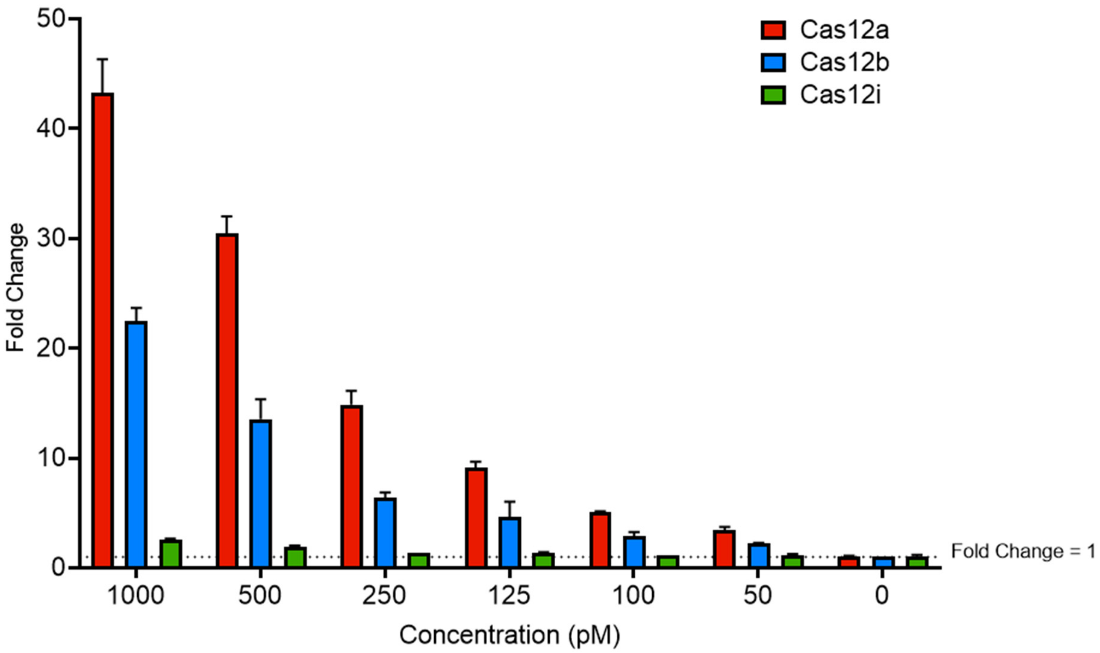


**Fig. S9:** Comparison of trans-cleavage activities of LbCas12a, BrCas12b, and Cas12i1. Figure showing the trans-cleavage activity of Cas12a, Cas12b, and Cas12i with target concentration varying from 1,000 to 0 pM. Here Cas12a is labeled red, Cas12b is labeled blue, and Cas12i is labeled green. Error bars represent Mean ± SD (n=3).

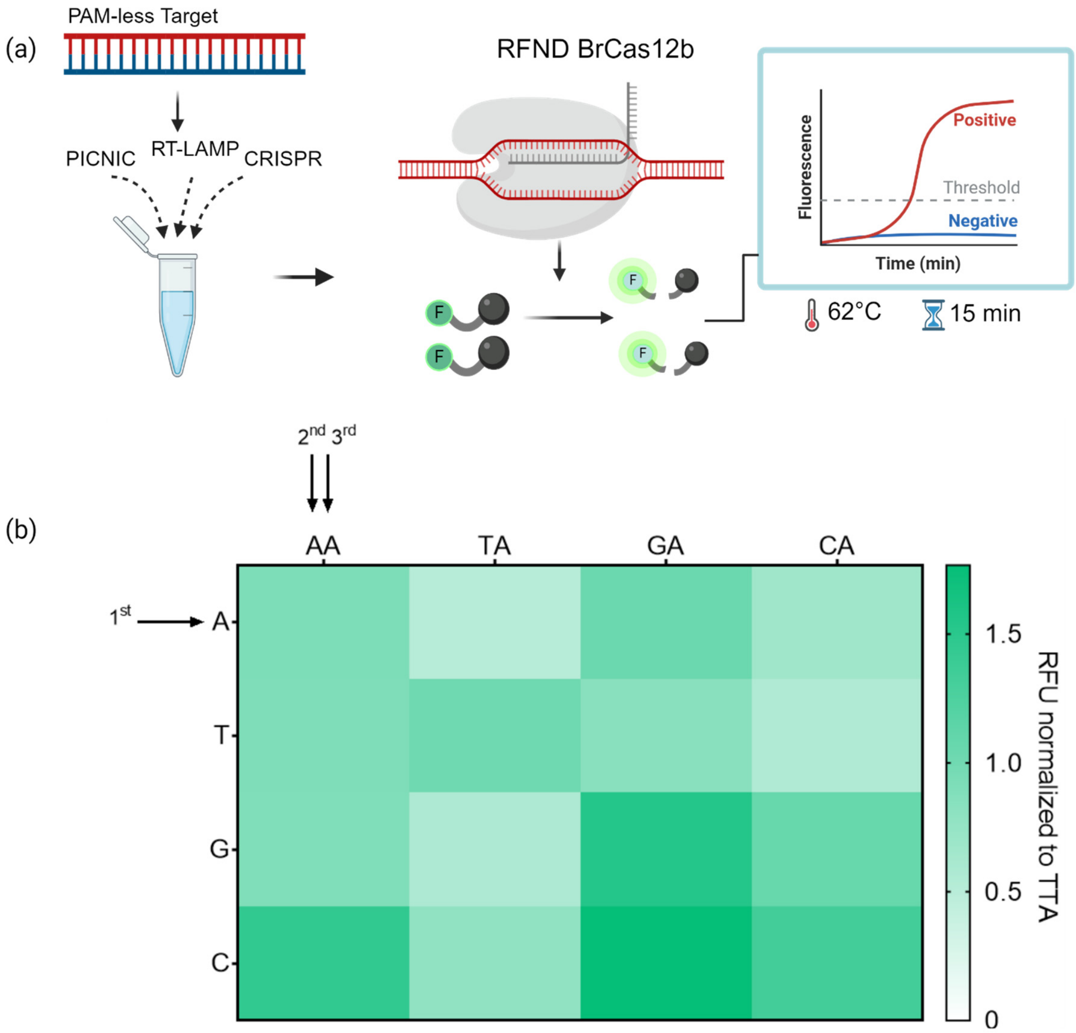


**Fig. S10:** Heat map represents the fold change of trans-cleavage of 15-nt truncated crRNAs containing A, U, G, or C bases at the first position, tested for the detection of targets containing T, A, C, or G base in a combinatorial fashion (n=3). The fold change is normalized to the activity of the canonical crRNA-target combination.

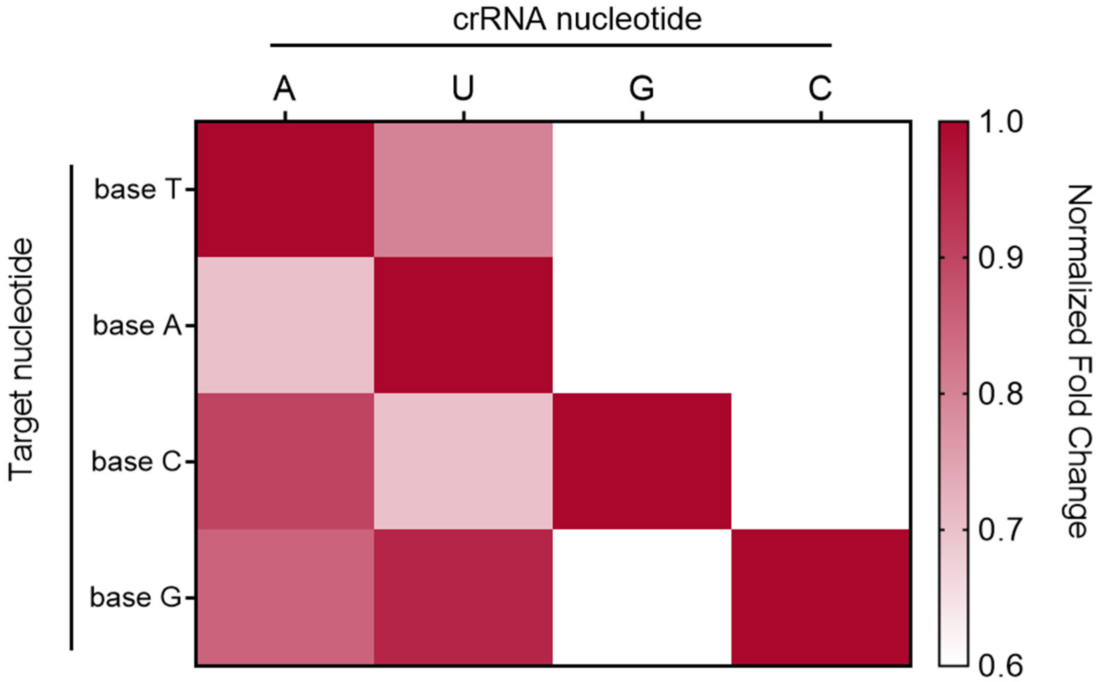


**Fig. S11:** Heat map represents the fold change of trans-cleavage of 15-nt truncated crRNAs containing A, U, G, or C bases at the first position, tested for the detection of targets containing T, A, C, or G base in a combinatorial fashion (n=3). The fold change is normalized to the activity of the canonical crRNA-target combination.

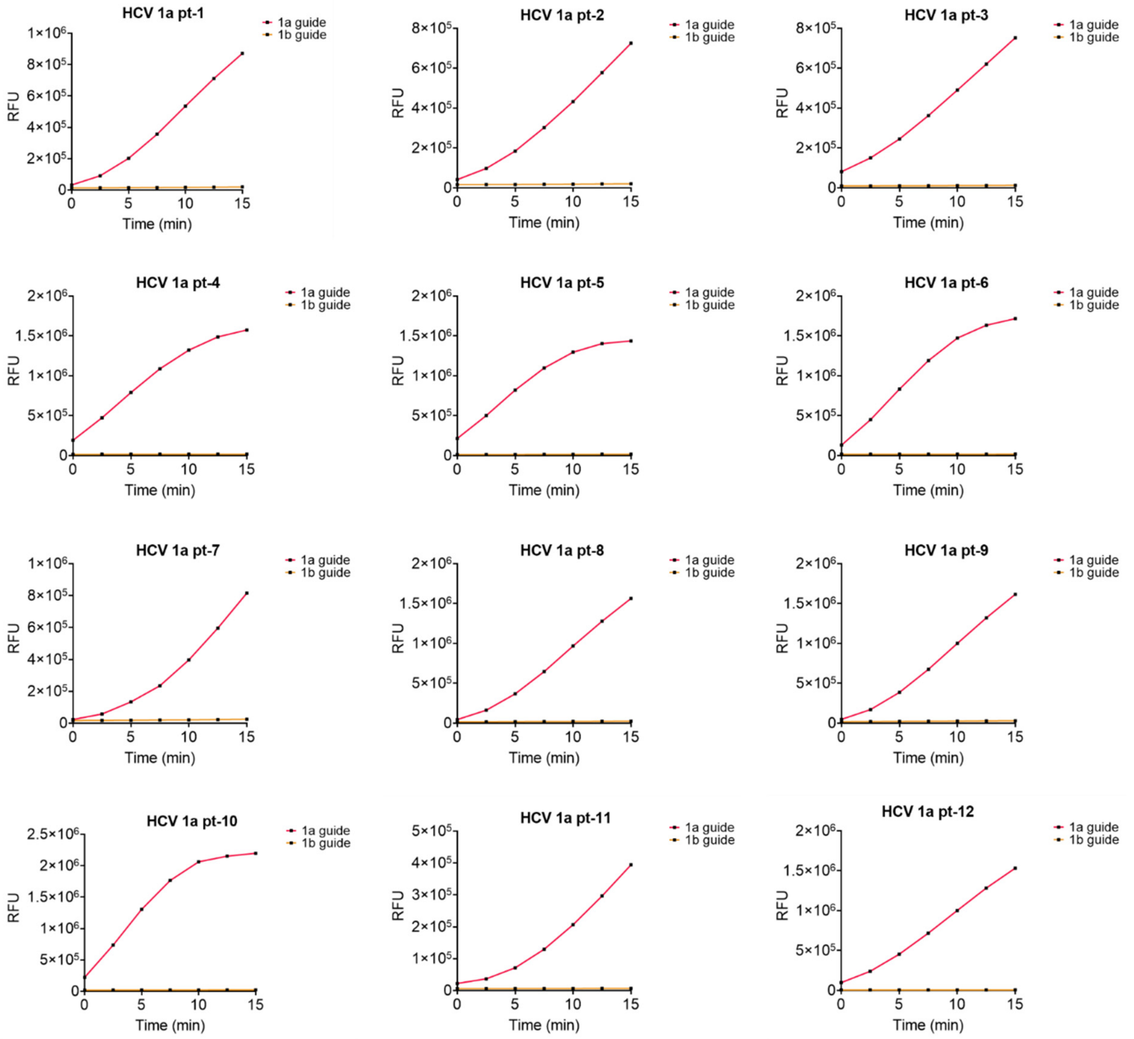


**Fig. S12:** Plot representing the intensity of the fluorescence obtained in RFU for the detection of 48 HCV-1a patient samples with a PICNIC-based test designed to genotype using specific crRNAs named HCV-1a guide RNA (red) and HCV-1b guide RNA (orange). The HCV-1a samples only show strong fluorescence in the presence of HCV-1a guide but not the HCV-1b guide.

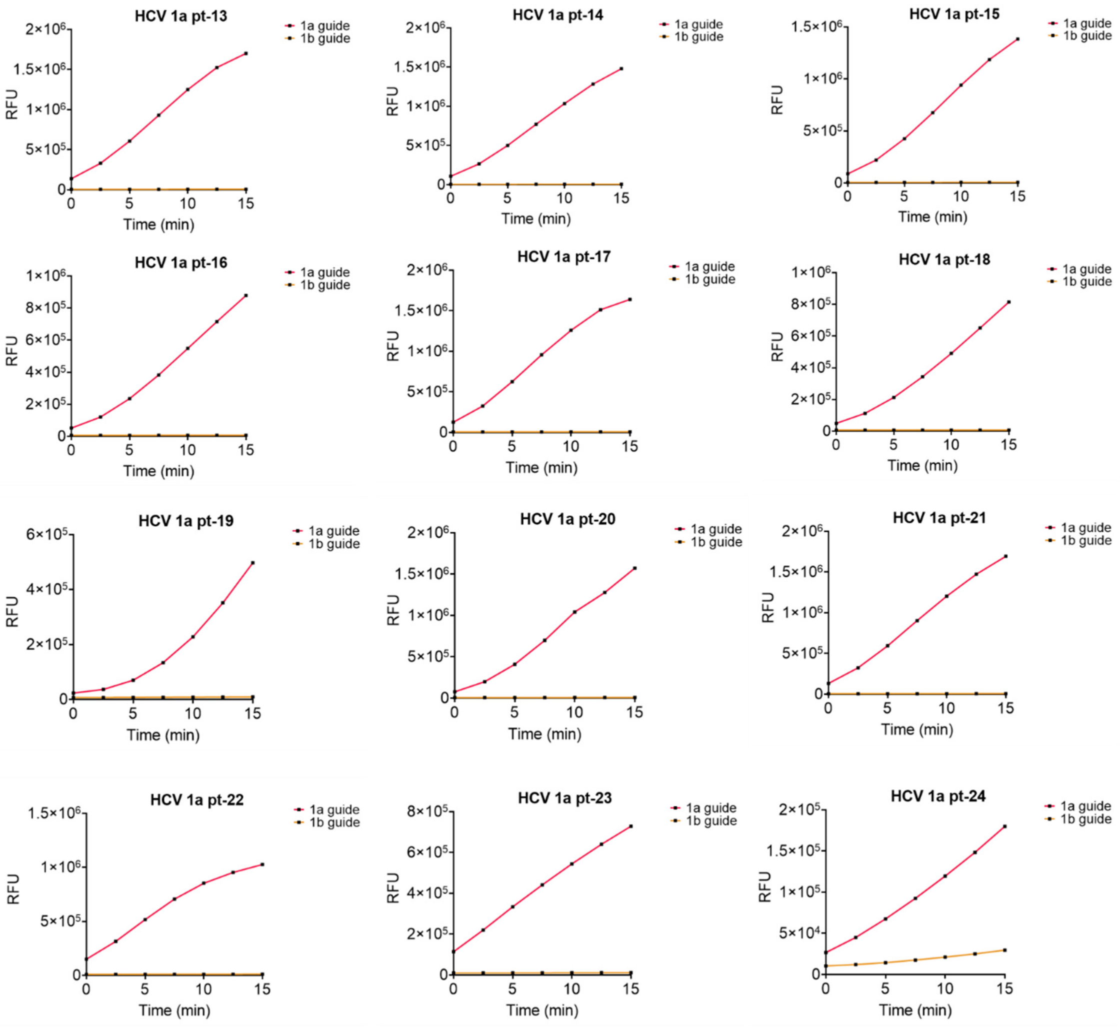


**Fig. S12 continued**

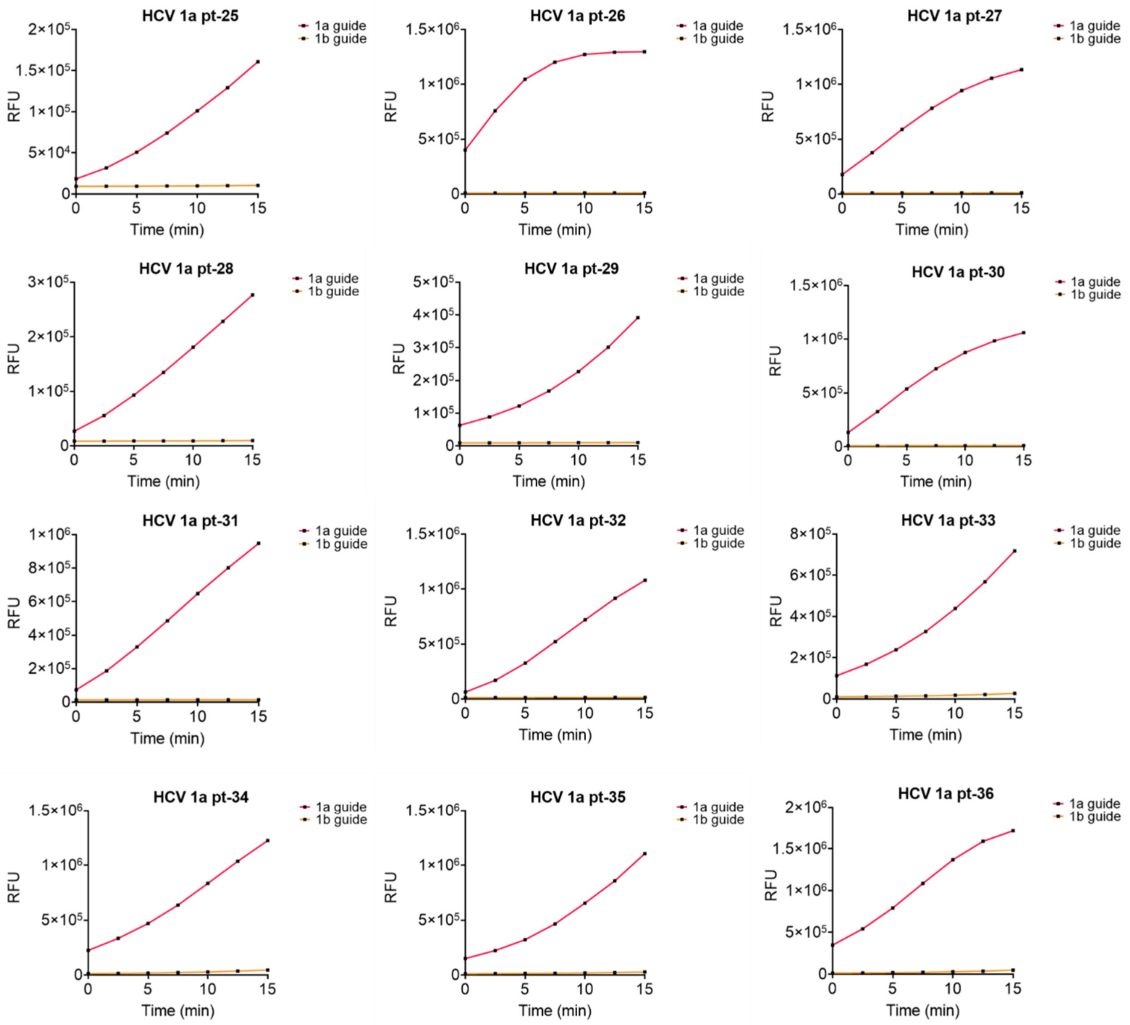


**Fig. S12 continued**

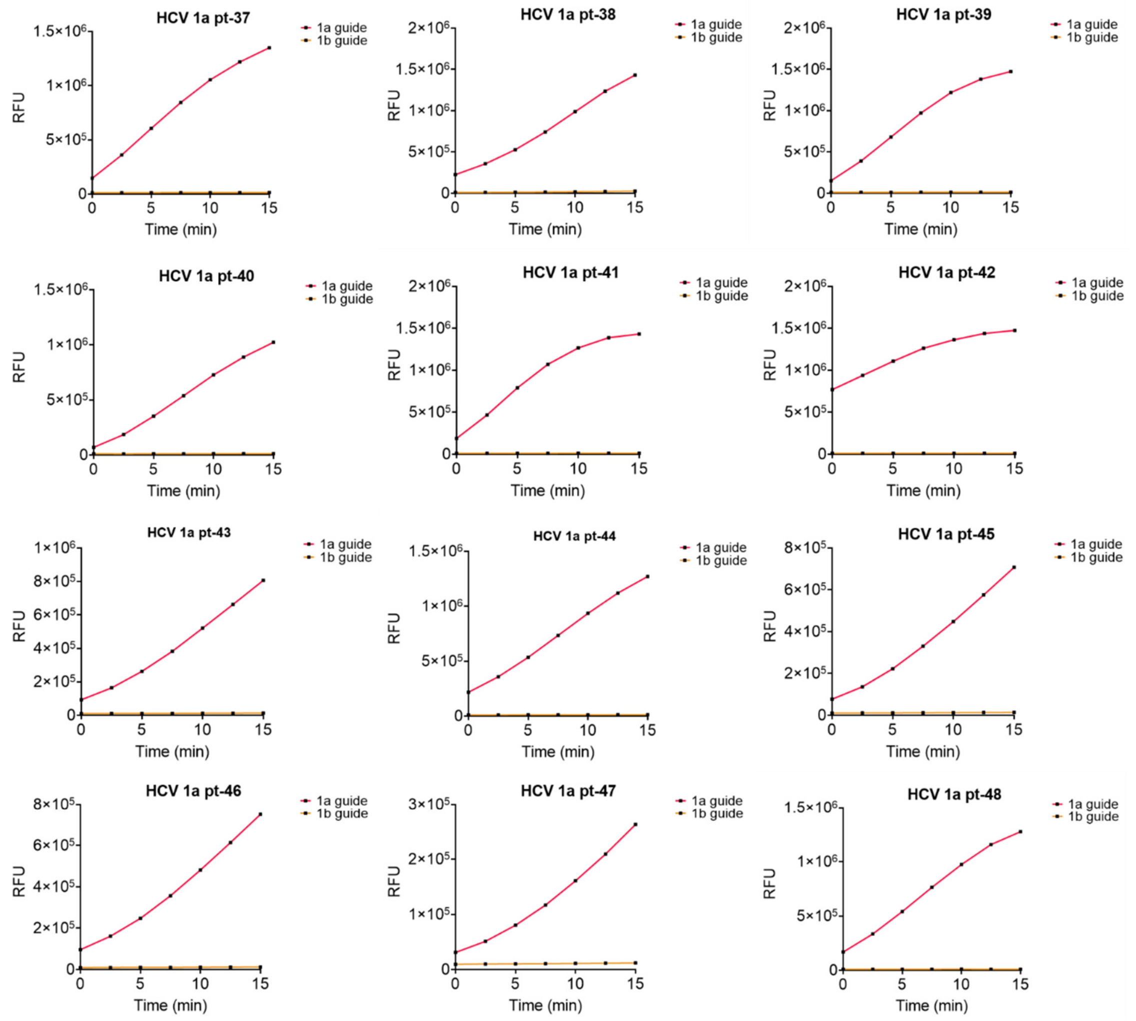


**Fig. S12 continued**

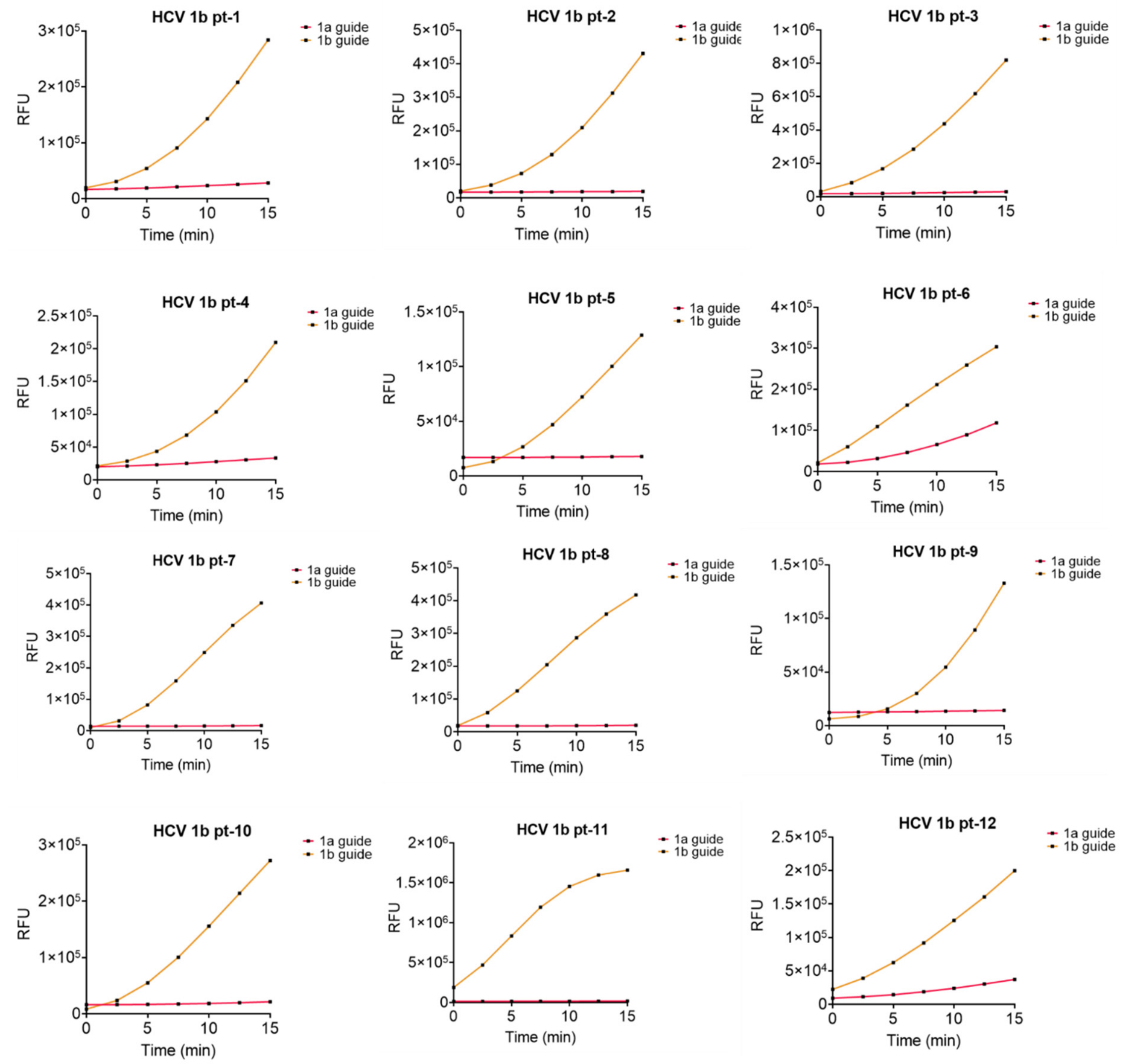


**Fig. S13:** Plot representing the intensity of the fluorescence obtained in RFU for the detection of 40 HCV-1b patient samples with a PICNIC-based test designed to genotype using specific crRNAs named HCV-1a guide RNA (red) and HCV-1b guide RNA (orange). The HCV-1b samples only show strong fluorescence in the presence of HCV-1b guide but not the HCV-1a guide.

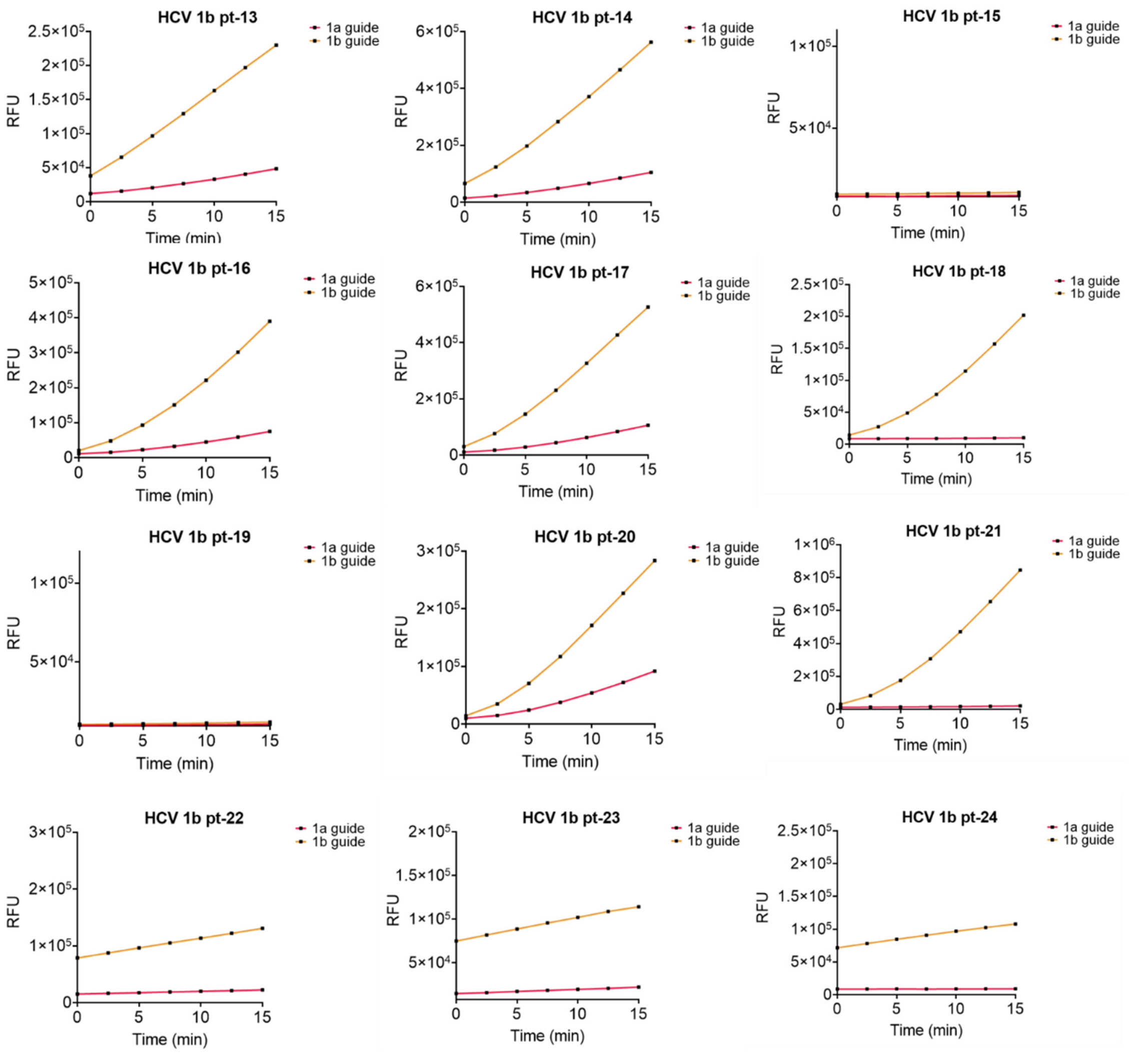


**Fig. S13 continued**

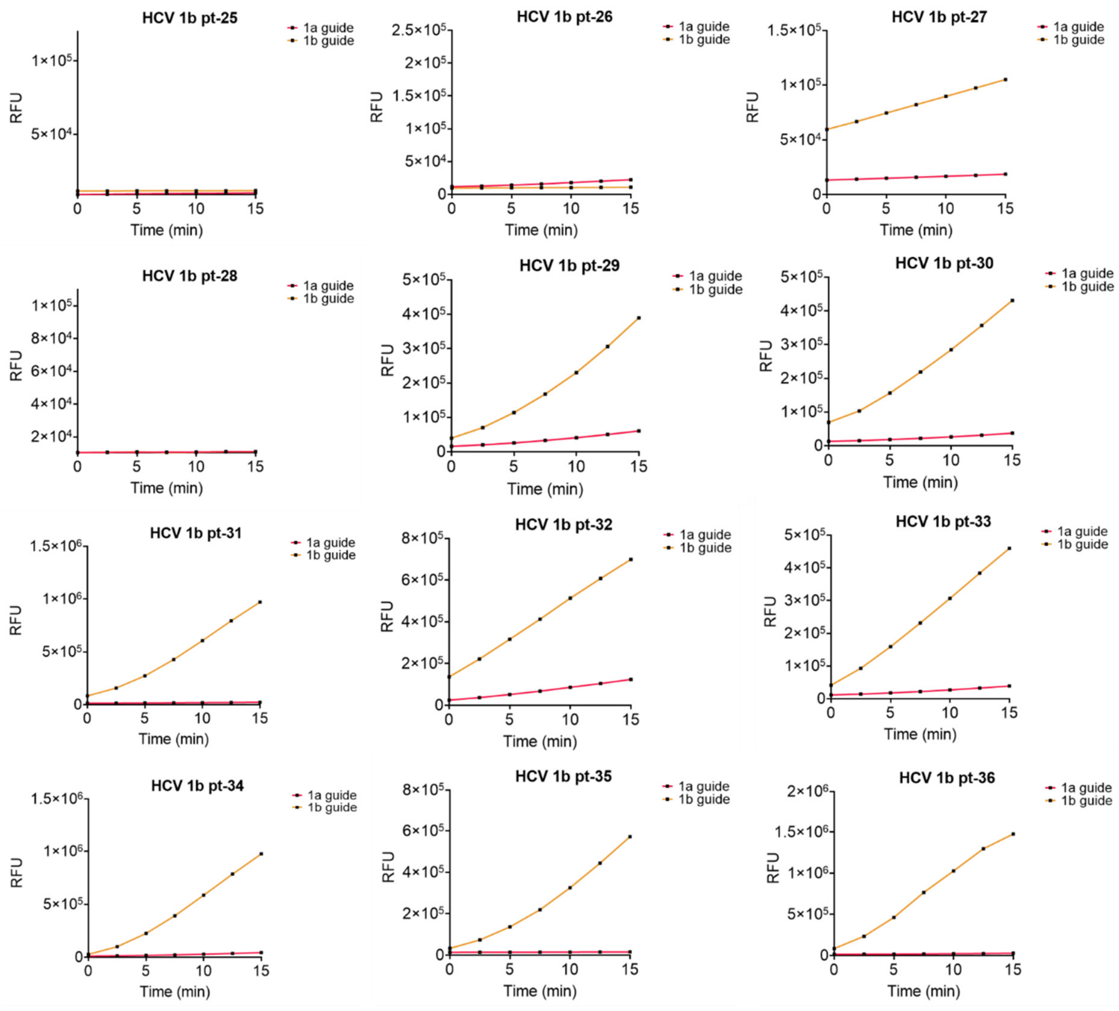


**Fig. S13 continued**

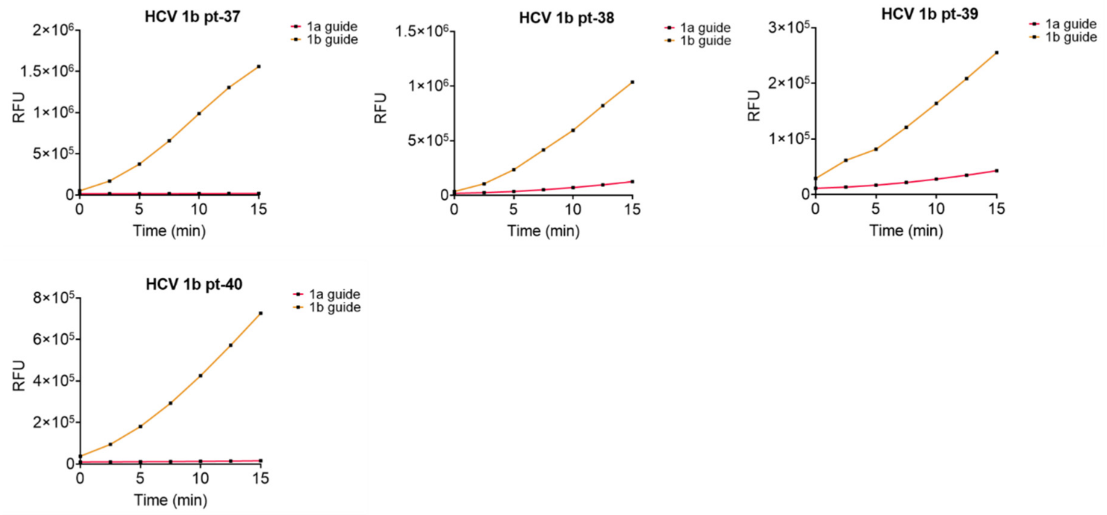


**Fig. S13 continued**
